# Heterozygous *KRT32* variant is responsible for autosomal dominant loose anagen hair syndrome

**DOI:** 10.1101/2025.05.02.25326729

**Authors:** Marcelo Melo, Elizabeth Phillippi, Thomas Moninger, Kya Foxx, Benjamin Darbro, Kelly N. Messingham, Edward A. Sander, Hatem El-Shanti

**Affiliations:** Department of Pediatrics, Carver College of Medicine, University of Iowa, Iowa City, IA 52242, USA; University of Iowa Central Microscopy Research Facility; MSTP summer program, Carver College of Medicine, University of Iowa, Iowa City, IA 52242, USA; Department of Dermatology, Carver College of Medicine, University of Iowa, Iowa City, IA 52242, USA; Roy J. Carver Department of Biomedical Engineering, University of Iowa, Iowa City, IA 52242, USA

**Keywords:** Loose Anagen Hair Syndrome, far western blot, fluorescent microscopy, intermediate filaments, KRT32, KRT82

## Abstract

Loose Anagen Hair Syndrome is a form of non-scarring alopecia marked by easily and painlessly pluckable terminal hair during its active growth – anagen – phase. This condition is believed to result from poor hair shaft anchoring within the follicle due to premature keratinization. Our research identified the likely pathogenic c.296C>T (p.T99I) variant in *KRT32*, which was found to co-segregate with the disorder in a large family with autosomal dominant loose anagen hair syndrome. This study aimed to explore the role of KRT32, previously unassociated with loose anagen hair, in hair anchorage and assess the functional impact of the p.T99I variant. We hypothesized that the p.T99I variant reduces KRT32’s binding affinity to KRT82, disrupting the intermediate filament structure in the hair shaft cuticle and leading to weak anagen hair anchorage. To test this hypothesis, we conducted a protein-protein interaction assay using far western blotting and performed *in silico* intermediate filament network segmentation analysis on high-resolution fluorescent microscopy images. Our results revealed a decreased binding affinity of KRT32^T99I^ for KRT82 compared to KRT32^WT^, along with significant differences in segment count and filament brightness (thickness) between the two groups.

## 1. Introduction

Loose anagen hair syndrome (LAHS) is a rare form of non-scarring alopecia characterized by poorly anchored hair shafts during the growth (anagen) phase ^1, 2^. It is associated with hair thinning that is usually diffuse, but can be patchy; the anagen hairs can be easily and painlessly plucked and are lost without proceeding to the catagen then telogen phases manifesting as hair that seemingly does not grow ^3, 4^. The diagnosis of LAHS is achieved by the presence of a positive hair-pull test in which most of the pulled hair is in the anagen phase ^3^. On microscopic examination, the extracted hairs lack the inner and outer root sheaths (IRS and ORS) and show misshapen bulbs and ruffled or peeling cuticle layer proximal to the root ^2, 4^. Additionally, thin sections of scalp tissue containing affected hair follicles show disorganized keratin fibers, enlarged or distorted cells of the three IRS layers, and edema between the anchoring layers of the Henle and Huxley cells that help bind a normal hair shaft to the follicle ^5^.

Hair loss disorders that occur in the pediatric age group, including LAHS and another overlapping condition, short anagen hair syndrome (SAHS), are associated with bullying, decreased self-esteem, and reduced quality of life ^6, 7^. About half of LAHS and SAHS patients and their caregivers report negative psychological symptoms in the form of anxiety, depression, low self-esteem, and body dysmorphia, and about a third report bullying or mistreatment by their peers ^7^. The psychological impact of LAHS and SAHS is comparable to the impact of alopecia areata on children ^6, 7^.

LAHS can occur alone, may be part of a complex phenotype, such as Noonan ^8^ and tricho-rhino-phalangeal ^9^ syndromes, or may accompany other hair shaft pathologies, such as wooly hair or unmanageable hair ^10-12^, thus exhibiting phenotypic heterogeneity. LAHS occurs sporadically, but can be familial with an autosomal dominant pattern of inheritance ^13^. It is suggested that premature or defective keratinization occurs in the IRS cells which decreases the ability of the hair follicle to hold on to the growing hair shaft ^2^. One study suggested that LAHS is a keratin disorder, and identified a pathogenic variant in *KRT75* in three out of the nine studied families ^13^, however, this has not been replicated. Keratin 75 is expressed in the companion layer, previously considered the innermost layer of the outer root sheath (ORS). A recent study of hypotrichosis with loose anagen hairs identified compound heterozygous missense variants in *TKFC* in 1 out of 15 studied families and provided experimental functional evidence of the pathogenicity of the two variants ^14^. However, biallelic variants in *TKFC* are implicated in other multisystem phenotypes that do not include hair abnormalities ^15^.

We describe a three-generation family with multiple individuals affected by LAHS that fits an autosomal dominant pattern of inheritance. We identified a missense variant in *KRT32* that segregates with the phenotype. We provide in vitro experimental evidence that the missense *KRT32* variant reduces the binding affinity of Keratin32 to its partner, Keratin82, and reduces the number and thickness of the intermediate filaments. We anticipate that alteration in the intermediate filament structure affects the shape of the cells of the hair shaft cuticle decreasing the mechanical interlocking between the cuticle layers of the IRS and the hair shaft.

## 2. Material and Methods

### 2.1 Clinical Information

We obtained a release of information to acquire the medical records of the affected individuals, including the hair-pull test and microscopic examination of the extracted hairs when available. We constructed the pedigree (Figure 1 A).

**Figure 1.**
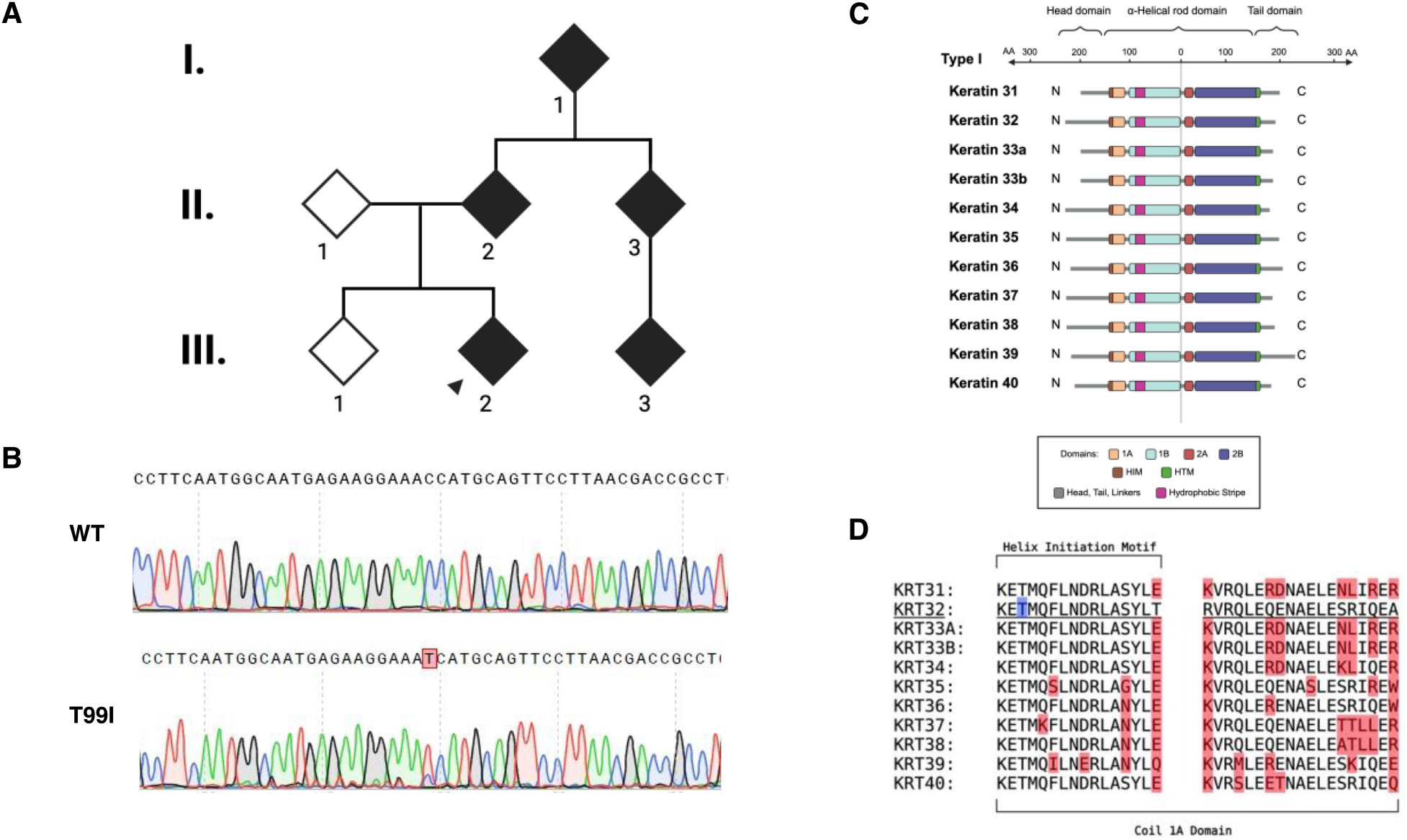
(A) Partial pedigree for the family studied. Symbols filled in black indicate individuals affected by LAHS. (B) Sanger sequencing chromatograph for unaffected (WT) and affected (T99I) individuals, with the site of the missense variant highlighted in red. (C) Diagram of all type I hair keratins, showing to-scale mapping of the amino acid length of various domains. Specific structural elements including protein domains, hydrophobic stripe, Helix Initiation Motif (HIM) and Helix Termination Motif (HTM), are color-coded, as annotated in figure key. (D) Amino acid sequence for coil 1A domain and HIM of type I hair keratins. Sequence for KRT32 is underlined, with p.T99I highlighted in blue. Red highlights amino acids that differ positionally from the KRT32 sequence (sequences obtained from UniProt). (Created with BioRender.com)

### 2.2 Biologic Samples

We obtained saliva samples (OGR-600, DNAgenotek, Inc., Ontario, Canada) for DNA extraction from all participating family members, after obtaining informed consent from the participants or their authorized legal representative.

### 2.3 Next-Generation Sequencing (NGS) and Variant Annotation

We performed genome sequencing (GS) for the proband, sibling, parents, and grandparent. Massively parallel GS was performed using the NovaSeq6000 platform (Illumina®, San Diego, CA, USA) and KAPA Hyper Prep Kit (Roche, Indianapolis, IN, USA) for 300 cycles at the Iowa Institute of Human Genetics (Iowa City, IA, USA) and at the S.R.P Molecular and Cytogenetics Laboratory at the University of Iowa. The paired-end reads were analyzed using the DRAGEN platform for read alignment, and variant calling (Illumina®, San Diego, CA, USA). Variants were annotated using the VarSeq platform (Golden Helix, Bozeman, MT, USA). Variants present in the proband were then filtered using the following parameters: quality score ≥ 25, depth ≥ 8x, alternative allele frequency ≤ .002 or missing from: UK10K, 1KG, gnomAD, and Kaviar. Variants were further considered if their CADD score ≥ 15 or SpliceAI calculated a delta score for any category ≥ 0.2. missing from UK10K (https://www.uk10k.org/), 1000 genomes/1 KG (https://www.internationalgenome.org/) ^16^, gnomAD v.2.1(https://gnomad.broadinstitute.org/), and Kaviar (https://db.systemsbiology.net/kaviar/, accessed on 14 April 2025) ^17^. Filtered variants were further considered if their CADD score was ≥15 ^18^ or SpliceAI calculated a delta score for any category ≥ 0.2 ^19^. Appropriately rare and potentially pathogenic variants in the proband, their affected parent, and grandparent were then checked for segregation with the phenotype in the sequenced individuals. Variants were genotyped by Sanger sequencing in other family members to check segregation with the phenotype. The variants were then filtered biologically based on their role in hair follicle biology and hair shaft structure, hair follicle morphogenesis and cornification, signaling pathways for hair cycling, mechano-transduction, and extracellular matrix integrity and remodeling.

### 2.4 Cell Culture and Transfections

Human embryonic kidney cells (HEK 293) and immortalized human keratinocytes cells (HaCaT) were both used in the study. Both cell lines underwent mycoplasma testing and were authenticated and validated using STR analysis (ATCC, Manassas, VA, USA). Cells were cultured in Dulbecco’s Modified Eagle Medium (DMEM), supplemented with penicillin, streptomycin, and 10% fetal bovine serum (FBS) at 37°C and 5% CO_2_. HEK293 were cultured in Falcon^®^ 100mm TC-treated cell culture dishes (Corning Inc., Corning, NY, USA), and HaCaT cells in Falcon^®^ 4-well culture slides (Corning Inc., Corning, NY, USA). All expression plasmids used (c-Myc-KRT32^WT^, c-Myc-KRT32^T99I^, HA-KRT82^WT^, GFP-KRT32^WT^, GFP-KRT32^T99I^ and RFP-KRT82^WT^) were designed and ordered from GenScript (GenScript Biotech, Piscataway, NJ, USA). Transfections were performed using Lipofectamine 2000 (Thermo Scientific, Waltham, MA, USA) in Opti-MEM™ (Thermo Scientific, Waltham, MA, USA) following the protocol provided.

### 2.5 Cell Lysis and Protein Purification

HEK293 cells transfected with the c-Myc and HA-tagged constructs were lysed in a lysis buffer composed of 10mL RIPA Buffer (Alpha Teknova, Inc., Hollister, CA, USA) and 1 tablet of cOmplete™ Mini, EDTA-free Protease Inhibitor Cocktail (Roche Holding AG, Basel, Switzerland) using a sonicator. Tagged protein purification was performed using Pierce™ Anti-c-Myc magnetic beads (Thermo Scientific, Waltham, MA, USA) and Pierce™ Anti-HA magnetic beads (Thermo Scientific, Waltham, MA, USA) following the provided protocols. Purified proteins were quantified using the NanoOrange™ Protein Quantification Kit (Invitrogen, Waltham, MA, USA), and fluorescence measured using the TECAN infinite M200 PRO (Tecan, Männedorf, Switzerland).

### 2.6 Western Blotting

100ng of each purified c-Myc-tagged protein construct was incubated at 95°C for 5 minutes in 20μL of loading buffer. Loading buffer was made of 9 parts 4X NuPAGE™ LDS Sample Buffer (Invitrogen, Waltham, MA, USA) and 1 part Bond-Breaker™ TCEP Solution (Thermo Scientific, Waltham, MA, USA). Samples were then loaded into separate wells of a NuPAGE™ Bis-Tris Mini Protein Gel, 4-12% (Invitrogen, Waltham, MA, USA). Gels were run for 20 minutes at 80V followed by 1h and 20 minutes at 125V, in 1X NuPAGE™ MOPS SDS Running Buffer (Invitrogen, Waltham, MA, USA) using the Invitrogen Mini Gel Tank. PageRuler™ Plus Prestained Protein Ladder (Thermo Scientific, Waltham, MA, USA) was used as the molecular weight ladder. Transfer to Immobilon^®^-P Membrane (PVDF) (Millipore, Burlington, MA, USA) was performed on a Trans-Blot^®^ SD Semi-Dry Transfer Cell (Bio-Rad, Hercules, CA, USA) for 2h using 1X transfer buffer (Tris-base, glycine, SDS). Membranes were then blocked for 1h at RT with agitation using 5% milk in TBST. Primary antibody incubation was carried out overnight with agitation at 4°C using c-Myc Polyclonal Antibody (1:500, Cat# A00172-40) (GenScript Biotech, Piscataway, NJ, USA). Secondary antibody incubation was performed at RT with agitation for 2h using Goat-Anti-Rabbit IgG (H+L)-HRP Conjugate (1:2000, Cat# 1706515) (Bio-Rad, Hercules, CA, USA). SuperSignal™ West Pico PLUS Chemiluminescent Substrate (Thermo Scientific, Waltham, MA, USA) was used before chemiluminescence development on the iBright™ (Invitrogen, Waltham, MA, USA).

### 2.7 Far Western Blotting

Imaged membranes from WB were stripped of secondary antibodies for 30 minutes at RT with agitation using Restore™ Western Blot Stripping Buffer (Thermo Scientific, Waltham, MA, USA). Complete stripping of secondary antibodies was verified by reincubation in SuperSignal™ West Pico PLUS Chemiluminescent Substrate followed by a 5 minute exposure on the iBright. Membrane-bound proteins were then denatured and renatured by incubation in a series of buffers with decreasing concentrations of guanidine HCl (buffer composition in Appendix A). Renatured proteins were incubated in binding buffer (Appendix A) which included 200ng of HA-tagged KRT82^WT^ for 4h at RT with agitation. Once binding buffer was washed off, membrane were blocked, incubated in primary antibody HA Tag Monoclonal Antibody (1:2500, Cat# 26183) (Invitrogen, Waltham, MA, USA), secondary antibody Goat Anti-Mouse IgG (H + L)-HRP Conjugate (1:2000, Cat# 1706516) (Bio-Rad, Hercules, CA, USA), chemiluminescence substrate applied and imaged on iBright, as described above.

In summary, a western blot (WB) was performed using purified protein, referred to as prey protein. For this experiment KRT32^WT^ and KRT32^T99I^ served as prey proteins. Once the WB was concluded and imaged, bands were quantified in ImageJ, which served as a loading control for the downstream FWB probing. After quantification, the membrane was stripped of secondary antibodies and the membrane-bound proteins reantured. This is a crucial step because it allows the prey proteins to refold and adopt their native conformation required for binding. Once the prey proteins were renatured, they were incubated in a binding buffer containing the binding candidate referred to as bait protein. The incubation in bait protein is followed by incubations in antibodies against the bait and imaging to assess KRT82 binding to membrane-bound KRT32^WT^ and KRT32^T99I 20^

### 2.8 Cell Fixation

Post-transfection and pre-fixation, HaCaT cells’ nuclei were stained with viability stain NucBlue™ (Invitrogen). Slides were rinsed in ice-cold DPBS Ca^2+^ Mg^2+^ (Gibco, Waltham, MA, USA) and fixed for 5 minutes in cold 1% PFA, followed by additional washes in DPBS Ca^2+^ Mg^2+^. Slides were then incubated with 0.5% Triton X-100 for 5 minutes at room temperature (RT). Triton X-100 was rinsed and slides blocked with 4% BSA for 10 minutes at RT. Blocking buffer was replaced with diluted nanobodies [ChromoTek GFP-Booster Alexa Fluor 488 (1:250, Cat# gb2AF488) (Proteintech Group Inc, Rosemont, IL, USA) and ChromoTek RFP-Booster Alexa Fluor 568 (1:200, Cat# rb2AF647) (Proteintech Group Inc, Rosemont, IL, USA) and incubated for 1 hour protected from light. Slides were then washed again in DPBS Ca^2+^ Mg^2+^ and a final time in DPBS -/- for 5 minutes. Slides were finally mounted with ProLong™ Diamond antifade (Invitrogen, Waltham, MA, USA), coverslipped, and stored protected from light at -20°C until imaging.

### 2.9 Microscopy

To obtain statistically relevant data (post-hoc power calculations Appendix B), more than forty image data sets for each co-transfection pairing (GFP-KRT32^WT^/RFP-KRT82 and GFP-KRT32^T99I^/RFP-KRT82) were obtained on a Zeiss LSM980 equipped with Airyscan 2 super-resolution detector (Zeiss, Oberkochen, Germany). Fluorescence probes were excited with 561 nm, 488 nm, and 405 nm lasers with detection bands set to BP 573-627, 495-550, and 422-477, respectively. Optimal Airyscan settings for z-stacks were used, resulting in a voxel size of xy = 0.035μm and z = 0.13μm. An oil immersion 63 x/1.4 object lens was used.

### 2.10 Image Processing and Analysis

Partially processed 4-channel “Sheppard Ring” Zeiss CZI Airyscan image files were deconvoluted using SVI Huygen’s Professional Deconvolution (Scientific Volume Imaging B.V., Hilversum, Netherlands). Considering the RFP and GFP fluorescent signals overlapped, for the remainder of the workflow only the GFP channel was used as it consistently generated the strongest fluorescent signal, facilitating the IF network mapping by TSOAX. No further image optimization was used.

## Results

### 3.1 Clinical history and phenotype

The proband, currently a teenager, has fine, short, sparse hair. They were diagnosed with LAHS as a child, because of the sparse hair that is easily extracted and by a positive hair-pull test that showed mainly anagen hairs with absent root sheaths, misshapen bulbs, and ruffled cuticles. The family history is consistent with penetrant autosomal dominant pattern of inheritance. Their symptoms improved after puberty, and with treatment with Minoxidil and the diagnosis of short anagen hair syndrome appeared in their medical records. One parent, II.2, has a history of sparse hair that improved around puberty, minimal body hair, brittle nails, and missing wisdom teeth. The post-pubertal improvement is mainly in the scalp hair, while body hair, eyebrows, and eyelashes remain sparse. Affected males, usually contend with early-onset baldness. The proband’s affected parent’s sibling, II.3, has a similar phenotype as well as their child, III.3, who is diagnosed with LAHS by a hair-pull test.

### 3.2 Variant identification and analysis

Our group identified a large 3-generational family with multiple individuals affected by LAHS with seemingly autosomal dominant inheritance pattern (Figure 1A). Genome sequencing analysis identified a missense heterozygous variant in *KRT32* (NM_002278.3:c.296C>T, p.Thr99Ile), present in the proband, parent, II.2 and grandparent, I.1, and absent in sibling, III.1, and parent, II.1. The variant has a minor allele frequency (MAF) of 0.00162 in the public access databases. Variant p.T99I was found to also be present in relatives II.3, and III.3, by Sanger sequencing, showing that it segregates with the phenotype and is inherited as an autosomal dominant trait. The variant p.T99I is predicted to be disease-causing by six *in silico* protein prediction models (SIFT, Polyphen2 HVAR, MutationTaster, MutationAssessor, FATHMM Pred, and FATHMM MKL Coding Prediction), likely by hindering its ability to efficiently bind with its type II counterpart, and has a CADD score of 20.6. The KRT82 encoded by *KRT82* is a type II hair keratin, reported to form IF in the cuticle layer of the hair shaft by heterodimerization with KRT32 ^21, 22^. The threonine residue in position 99 in KRT32 is highly conserved evolutionarily, not only in mammals, but also in chicken, *Xenopus tropicalis*, and zebrafish. The SWISS-MODEL and AlphaFold Server (Beta) did not predict any significant structural changes to KRT32 by introducing variant p.T99I. The variant p.T99I is in the N-terminus helix initiation motif (HIM) within the central α-helical rod domain of KRT32 (Figure 1C); HIM is a highly conserved region across all keratins (Figure 1D). Most pathogenic variants encountered in keratins are in the HIM or in the also highly conserved C-terminus helical termination motif (HTM) ^23^.

### 3.3 Protein-protein interaction assay

Since the *in silico* protein prediction models depicted that the variant may hinder the ability of KRT32 to efficiently bind with its type II counterpart, KRT82, we performed far western blots (FWB) to directly compare the binding affinity of KRT32^T99I^ and KRT32^WT^ to KRT82. FWB is a protein-protein interaction assay where binding affinity can be measured by quantifying band intensity on a membrane.

Four WB and FWB pairings were performed, and chemiluminescence absorbance was quantified as described in the methods section above. Absorbances of FWB bands were divided by the corresponding band from the WB, generating a binding coefficient for KRT82 and the respective prey protein construct. Combined analysis of all FWB quantifications shows an average 20.7% reduction in binding affinity of KRT32^T99I^ to KRT82 when compared to that of KRT32^WT^, with P=0.0329 (t-test) (Figure 2A-B). These findings support the hypothesis that variant p.T99I hinders the binding of KRT32 to KRT82.

**Figure 2.**
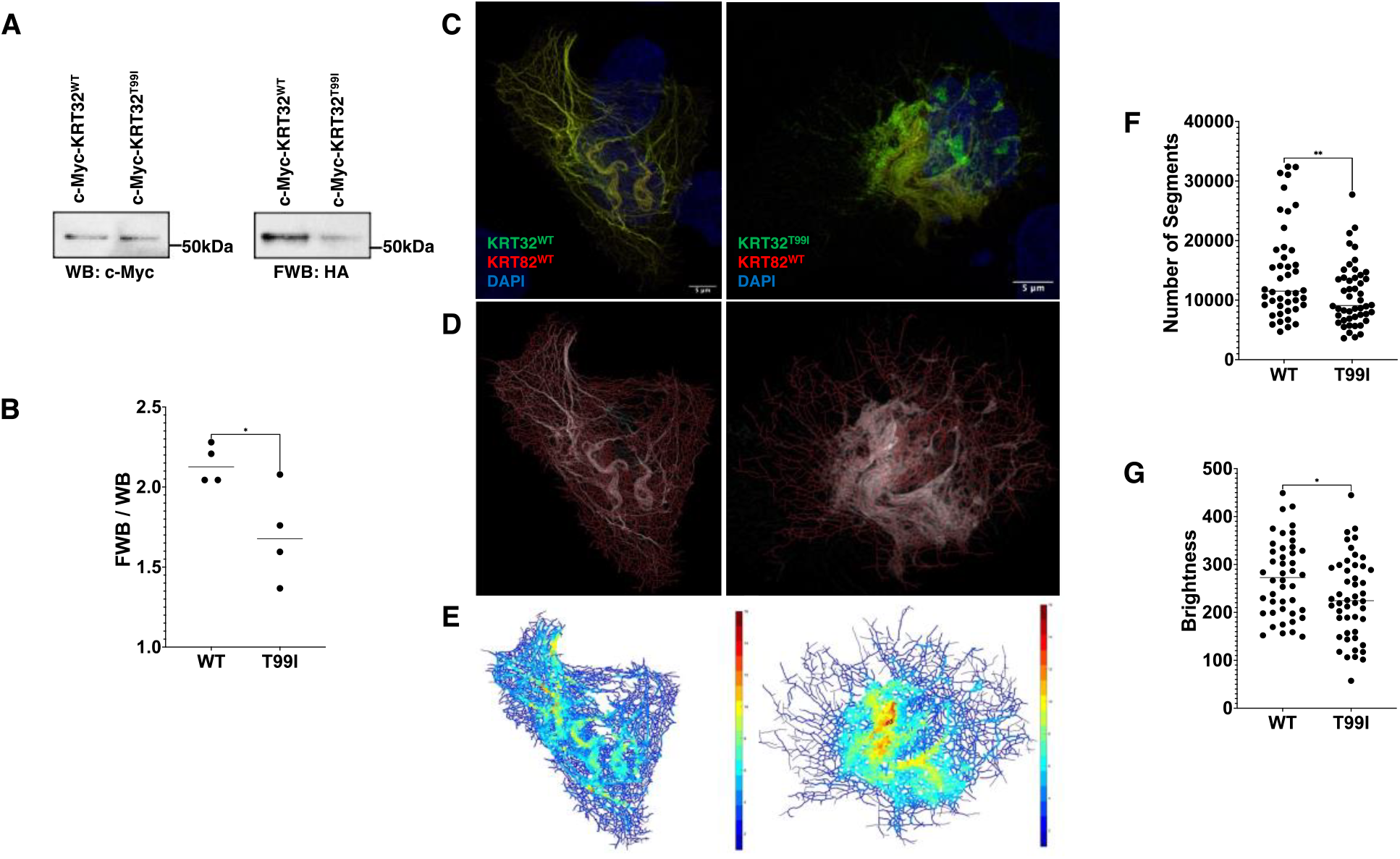
(A) Membrane images for WB (loading control) and FWB showing a difference between binding affinity between prey proteins KRT32^WT^ and KRT32^T99I^ and bait KRT82. (B) Summarized results for FWB experiments (n=4) showing binding affinity between prey proteins KRT32^WT^ and KRT32^T99I^ and bait KRT82, as a function of chemiluminescence absorbance (FWB/WB) P=0.0329 (t-test). (C) Maximum intensity projection of cells co-transfected with GFP-KRT32^WT^ or GFP-KRT32^T99I^ (green channel), RFP-KRT82^WT^ (red channel), and stained with DAPI (blue channel). (D) Overlay of the keratin network, generated by TSOAX, on their corresponding maximum intensity projections. (E) Keratin filaments color-coded by filament brightness which is reported to correspond to filament thickness. (F) Statistical analysis comparing the number of segments between the WT (n=44) and T99I (n=48) groups, showing a significant reduction for the T99I group P=0.0080 (t-test). (G) Statistical analysis comparing the average brightness between the WT (n=44) and T99I (n=48) groups, showing a significant reduction for the T99I group P=0.0151 (t-test).

### 3.4 Microscopy

To investigate the effect of the reduced binding affinity of KRT32^T99I^ to KRT82 has on IF organization and overall cell morphology, we performed image analysis of high-resolution fluorescence microscopy data of epithelial cells. For this, immortalized human keratinocytes HaCaT cells were co-transfected with RFP-tagged KRT82 and GFP-tagged KRT32^T99I^ expression plasmids and RFP-KRT82 with GFP-tagged KRT32^WT^ as a control. Keratin IF organization in cells was examined using Airyscan super-resolution microscopy (Figure 2C). IF analysis was done using open-source programs TSOAX ^24^, which enables the extraction of biopolymer filaments (segments) from image sets and the open-source code KerNet ^25^ which uses the nodal and vertices data from each segment to provide quantitative information on the properties of the keratin network and to facilitate 3D rendering of the network structure. The metrics analyzed included: the total number of segments in the networks, segment length, segment length-to-distance ratio (measure of segment straightness), curvature, brightness (assumed proportional to thickness), number of segments per node, segment angle, and variability of the segment angle from the principal direction of network orientation.

Figure 2D shows the TSOAX snake files overlayed the single-channel GFP maximum intensity projections from ImageJ (Rasband, W.S., U.S. National Institutes of Health, Maryland, USA) for each of the corresponding cells. The analysis shows a significant change in the total number of segments between the two groups with a P=0.0080 (Figure 2F). Average filament brightness (Figure 2E), reported as corresponding to filament thickness ^25^ was also significantly different (P=0.0151) for the two experimental groups (Figure 2G). All other metrics included in the analysis did not detect a statistically significant change between the groups. Image analysis of fluorescent microscopy data from HaCaT cells co-transfected with GFP and RFP-tagged plasmids showed a significant difference in the number and thickness (brightness) of filaments between the two groups. To ensure rigor and reproducibility, we selected 50 images from four independent transfection assays for each group. In addition, microscopy settings, including voxel size, were kept consistent across all data sets, and only validated and open-source codes were utilized in our analyses.

## 4. Discussion

LAHS is a non-scarring form of childhood-onset alopecia in which poorly anchored hair shafts can be easily and painlessly plucked ^1, 2^. While it seems to be a mild phenotype with low pathologic impact, especially that it improves with the onset of puberty, it still has a significant psychological impact on the affected children ^7^. LAHS exhibits remarkable phenotypic and genotypic heterogeneity, indicating the involvement of multiple genetic factors and different modes of inheritance patterns. Improvement in symptomatology is recognized and is probably influenced by endocrine, genetic, and environmental factors. The identification of the genetic factors contributing to the etiology of this rare syndrome is helpful in the understanding of the hair follicle pathophysiology and may provide information that is helpful with the classification and treatment of this and related disorders.

Our group studied a multi-generation family with multiple individuals with isolated LAHS that segregates in an autosomal dominant pattern and identified a missense variant in *KRT32* that segregates with the phenotype in the family, predicted to be disease-causing by *in silico* protein prediction algorithms, evolutionary conserved, and has a low MAF. *KRT32* is expressed in the cuticular cells of the hair shaft ^22^ and produces a protein that, together with KRT82, forms IF within these cells and is anticipated to contribute to their distinctive shape that interlocks with the cuticle of the IRS within the hair follicle ^21, 26^. We provide experimental evidence that the *KRT32* variant reduces the binding affinity of KRT32 to KRT82, producing fewer IF segments that are thinner than the wild-type IF. We now hypothesize that this reduced binding affinity and the production of fewer and thinner IF segments lead to morphological changes in the hair shaft and IRS cuticle layers that affect the interlocking mechanism, thus leading to poor mechanical anchoring of the hair shaft in its follicle.

Similar to other type I hair keratins, KRT32 will form heterodimers with other type II hair keratins ^27^. Keratins are, however, extremely promiscuous, with a type I being able to bind any type II ^28^. Among the known type II binding partners of KRT32 are KRT82 and KRT85, both expressed in the cuticle layer of the hair shaft ^22^. KRT82 is expressed in the mid-cuticle layer of the hair shaft, a region known to interlock with the cuticle of the IRS, providing mechanical anchorage ^21, 29^, whereas KRT85 is expressed in the bulb (lower portion) of the cuticle of the hair shaft ^22^. This observation supported our selection of KRT82 as a plausible partner of KRT32 in the functional studies performed in this study.

One limitation of this study was that all the functional experiments were conducted *in vitro* due to the absence of an appropriate animal model. Although mice have been utilized in hair loss research, they differ significantly from human terminal hair in both hair cycle regulation and morphology. Mouse hair is morphologically closer to vellus hair, found throughout the human body; it is lighter and shorter than the terminal hair found on the human scalp. The hair cycle in mice is synchronized between follicles, meaning they progress through phases of growth and shedding uniformly, whereas human hair follicles cycle asynchronously or independently from one another. Functionally, human terminal hair is also different than that of mice and other mammals. In mice, fur controls thermoregulation, whereas in humans, terminal hair protects the scalp tissue from ultraviolet radiation ^30, 31^. We propose using the recently developed human hair follicle organoid, generated entirely from pluripotent stem cells ^32, 33^ for further confirmation of the role of KRT32 and KRT82 in the anchoring of the hair shaft to its follicle and to provide an accurate model to identify other genes and study their effect on hair follicle biology.

## Data Availability

All data produced in the present study are available upon reasonable request to the authors

