## Appendix A for "Heterozygous *KRT32* variant is responsible for autosomal dominant loose anagen hair syndrome"

Protein-protein interaction assay

***Vectors***: designed and ordered from GenScript

- c-Myc-KRT32$^{WT}$

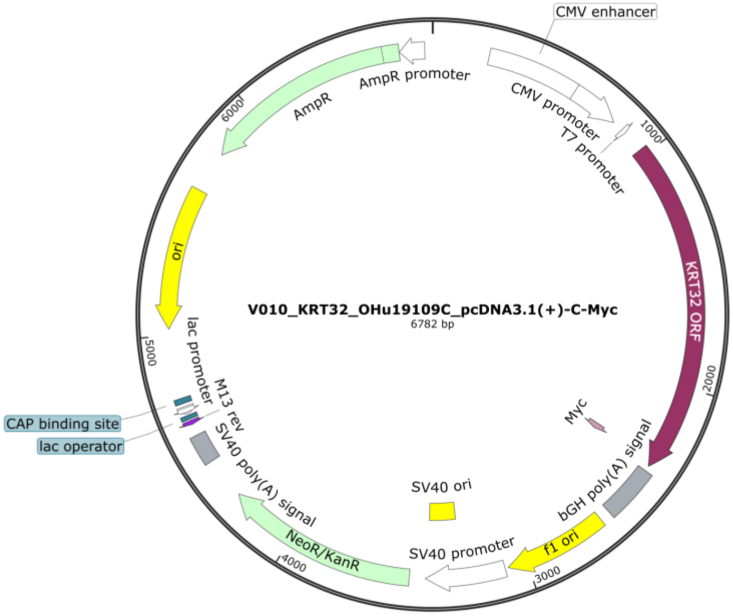

- c-Myc-KRT32$^{T99I}$

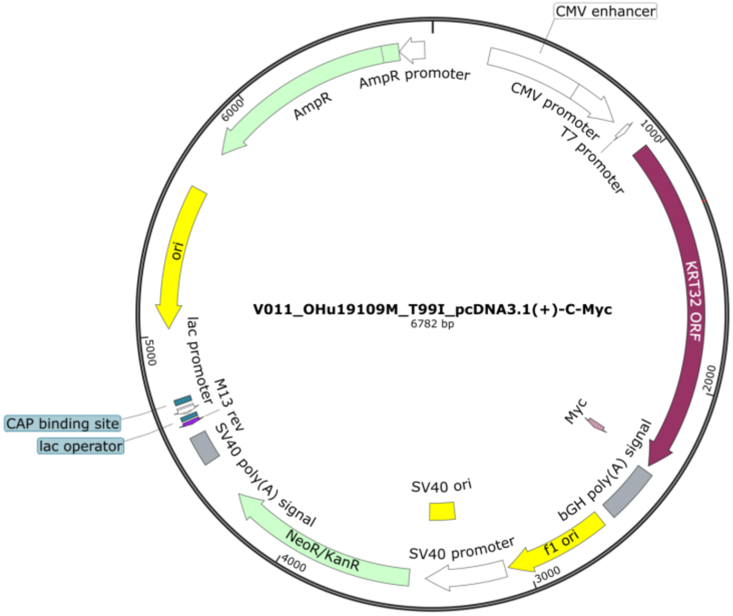

- HA-KRT82$^{WT}$

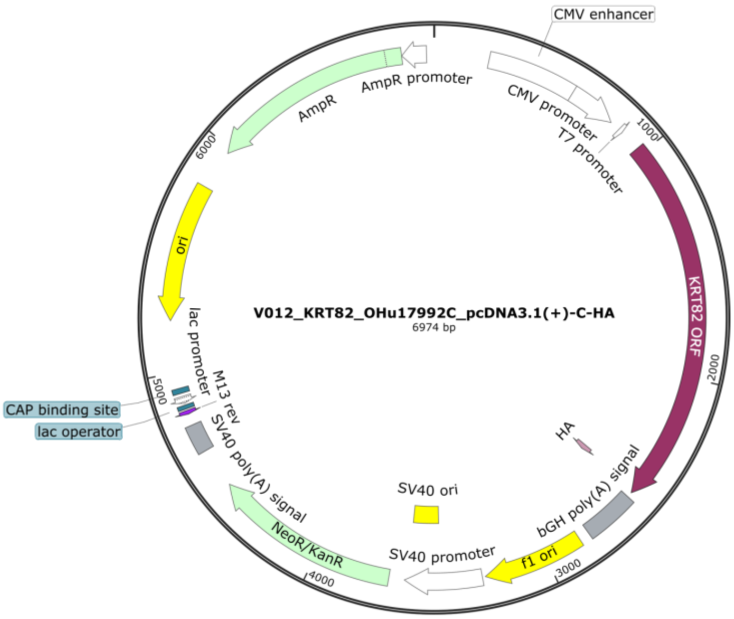

### ***Buffers:***

- Denature/Renature buffers recipe

| Concentration of guanidine-HCl (M) | 6 | 3 | 1 | 0.1 | 0 |
|---|---|---|---|---|---|
| Glycerol (ml) | 2.5 | 2.5 | 2.5 | 2.5 | 2.5 |
| 5 M NaCl (ml) | 0.5 | 0.5 | 0.5 | 0.5 | 0.5 |
| 1 M Tris, pH 8 (ml) | 0.5 | 0.5 | 0.5 | 0.5 | 0.5 |
| 0.5 M EDTA (ml) | 0.05 | 0.05 | 0.05 | 0.05 | 0.05 |
| 10% Tween-20 (ml) | 0.25 | 0.25 | 0.25 | 0.25 | 0.25 |
| Guanidine-HCl (8 M) (ml) | 18.75 | 9.30 | 3.13 | 0.31 | 0 |
| Milk powder (g) | 0.5 | 0.5 | 0.5 | 0.5 | 0.5 |
| 1 M DTT (μl) | 25 | 25 | 25 | 25 | 25 |
| ddH$_2$O (ml) | 2.45 | 12.82 | 18.07 | 20.89 | 21.20 |
| Total volume (ml) | 25 | 25 | 25 | 25 | 25 |
| Time/temperature | 30 min/room temperature (RT) | 30 min/RT | 30 min/RT | 30 min/4 °C | Overnight/4 °C |

- Protein-binding buffer composition

| Protein-binding buffer composition |
|---|
| 100 mM NaCl |
| 20 mM Tris, pH 8 |
| 0.5 mM EDTA |
| 10% Glycerol |
| 0.1% Tween-20 |
| 2% Milk powder |
| 1 mM DTT |

- Protein-binding buffer recipe (stock)

| Protein-binding buffer recipe (stock) | |
|---|---|
| 0.5 M EDTA (μl) | 50 |
| 1 M Tris, pH 8 (ml) | 1 |
| 5 M NaCl (ml) | 1 |
| 1 M DTT (μl) | 50 |
| $ddH_2O$ (ml) | 47.9 |
| Total volume (ml) | 50 |

- Reaction buffer recipe (working solution)

| 40 ml Reaction buffer (working solution) | |
|---|---|
| Glycerol (ml) | 4 |
| Milk powder (g) | 0.8 |
| Tween-20 (μl) | 40 |
| Protein-binding buffer – stock – (ml) | 36 |
| Total volume (ml) | 40 |

***Membrane:***

- Immobilon®-P Membrane (PVDF) Cat# IPVH85R Millipore

***Western Blot (WB) antibodies:***

- Primary: c-Myc Polyclonal Antibody (1:500, Cat# A00172-40) GenScript
- Secondary: Goat Anti-Rabbit IgG (H + L)-HRP Conjugate (1:2000, Cat# 1706515) Bio-Rad

***Stripping Buffer:***

- Restore™ Western Blot Stripping Buffer (Cat# 21059) Thermo Scientific™

***Far Western Blot (FWB) antibodies:***

- Primary: HA Tag Monoclonal Antibody (1:2500, Cat# 26183) Invitrogen
- Secondary: Goat Anti-Mouse IgG (H + L)-HRP Conjugate (1:2000, Cat# 1706516) Bio-Rad

***Cropped RAW membranes:***

- Experiment 1:

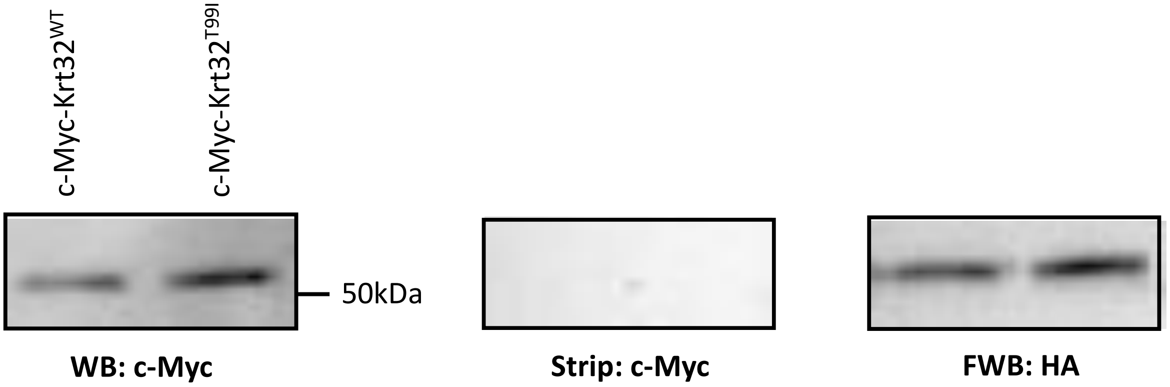

- Experiment 2: Bands from same membrane, imaged at the same time. Cropped and repositioned for parallel comparison.

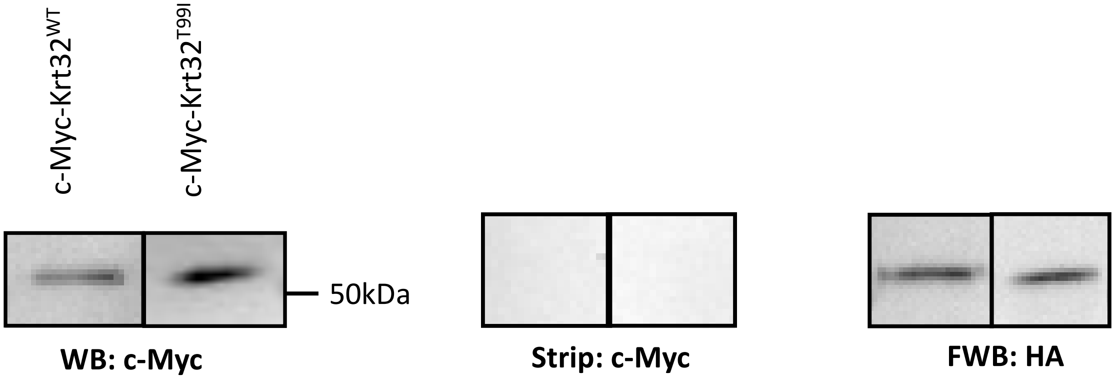

- Experiment 3:

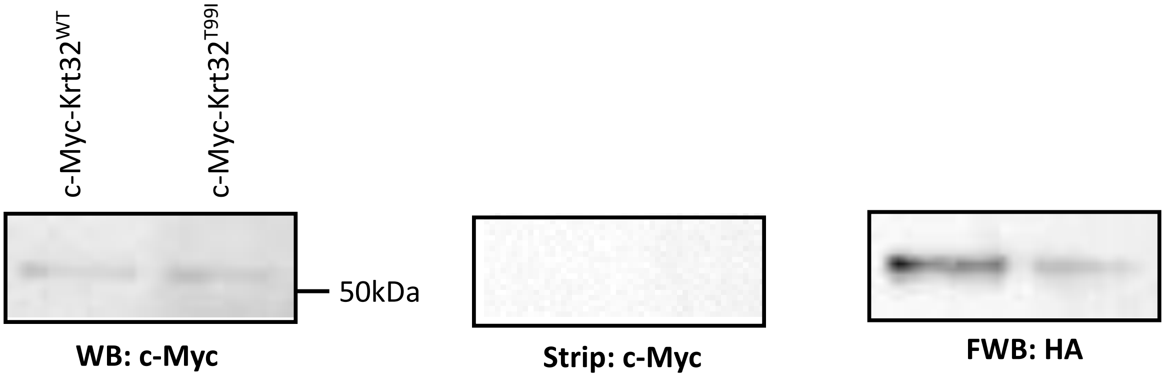

- Experiment 4:

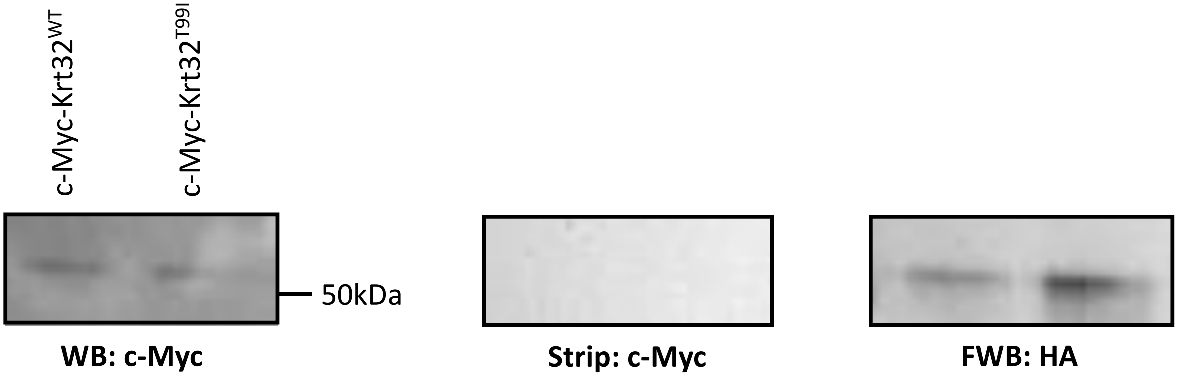

***Quantification and Statistical Analysis of Blots***

- Experiment 1:

| | KRT32$^{WT}$ | KRT32$^{T99I}$ |
|---|---|---|
| **WB** | 1417.92 | 1793.92 |
| **FWB** | 2898.406 | 3159.113 |
| **FWB/WB** | 2.044 | 1.76 |

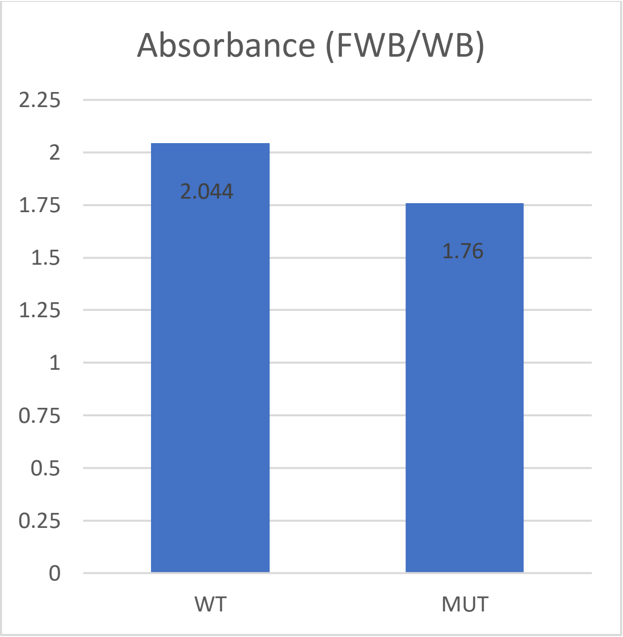

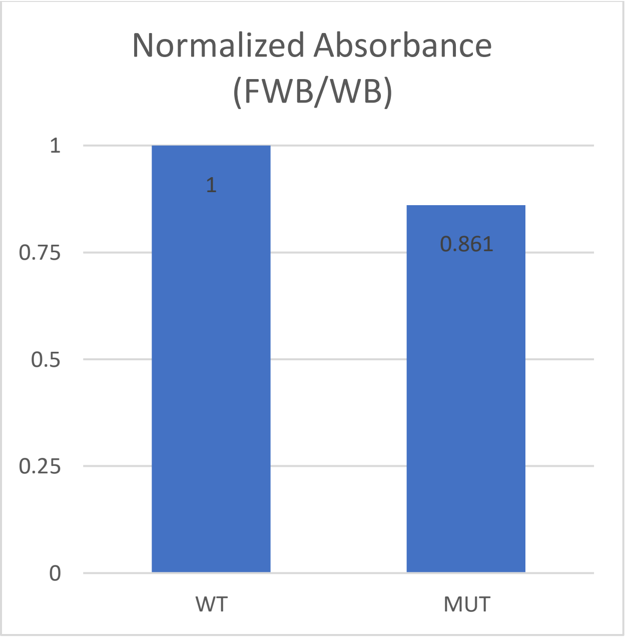

- Experiment 2:

| | KRT32$^{WT}$ | KRT32$^{T99I}$ |
|---|---|---|
| **WB** | 1417.92 | 1847.385 |
| **FWB** | 2898.406 | 2946.284 |
| **FWB/WB** | 2.044 | 1.595 |

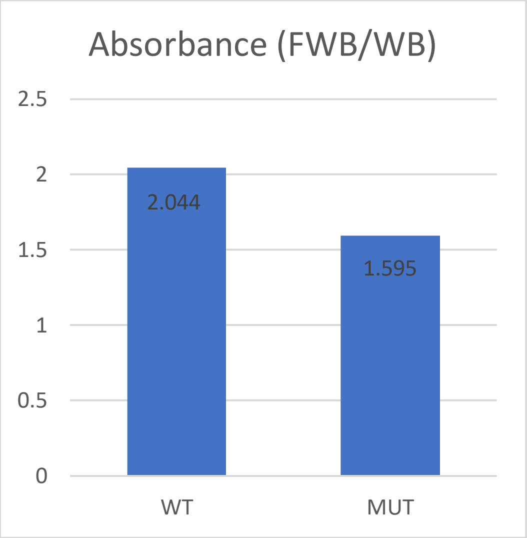

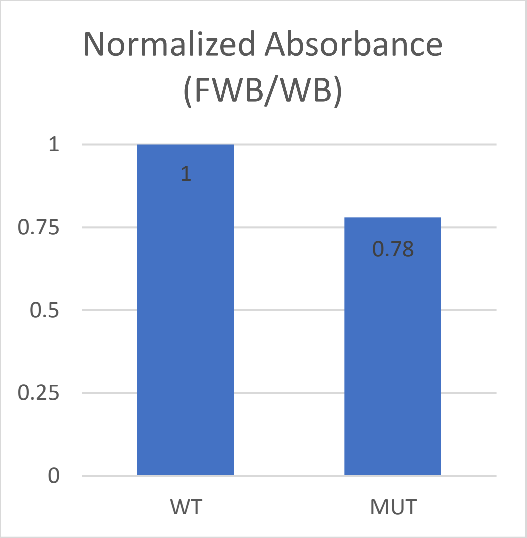

- Experiment 3:

| | KRT32$^{WT}$ | KRT32$^{T99I}$ |
|---|---|---|
| **WB** | 1607.799 | 809.728 |
| **FWB** | 3665.184 | 1107.113 |
| **FWB/WB** | 2.28 | 1.367 |

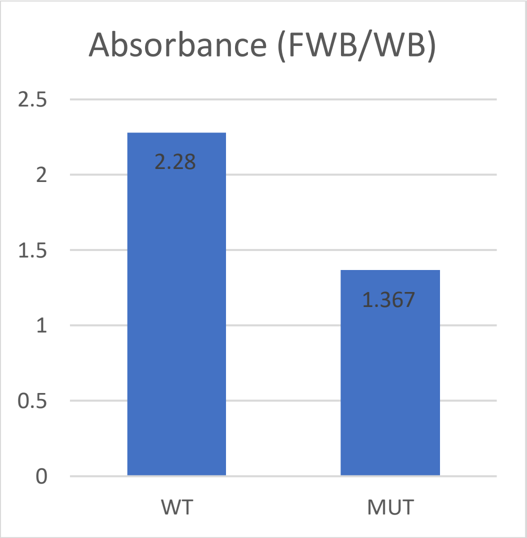

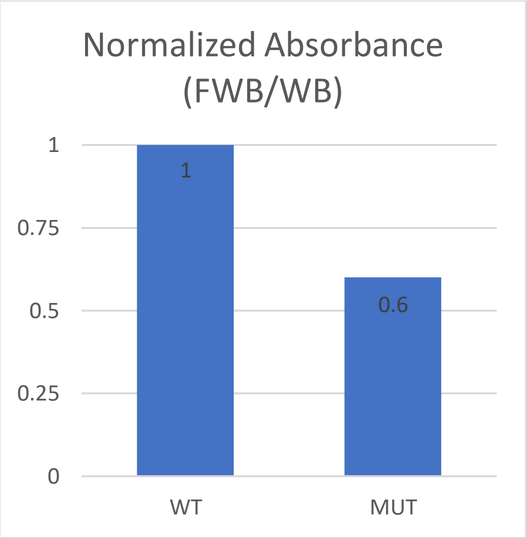

- Experiment 4:

| | KRT32$^{WT}$ | KRT32$^{T99I}$ |
|---|---|---|
| **WB** | 749.314 | 986.728 |
| **FWB** | 1654.577 | 2050.335 |
| **FWB/WB** | 2.208 | 2.078 |

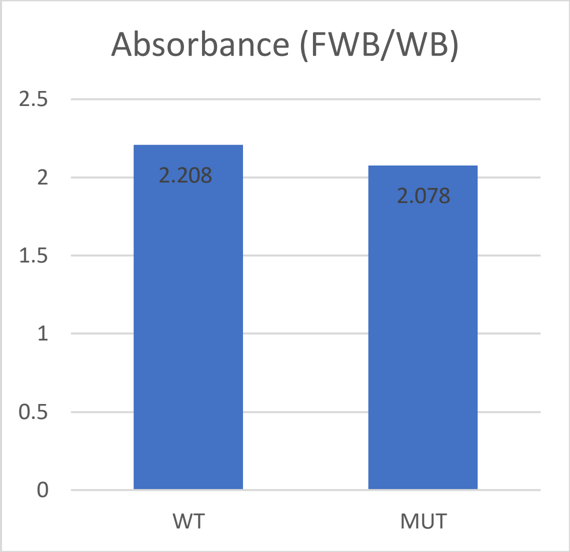

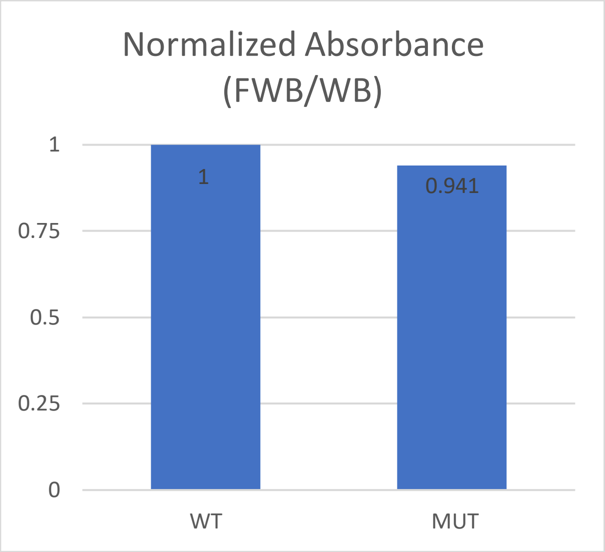
