## Appendix B for "Heterozygous *KRT32* variant is responsible for autosomal dominant loose anagen hair syndrome"

### Microscopy

***Vectors*:** designed and ordered from GenScript

- GFP-KRT32^WT^

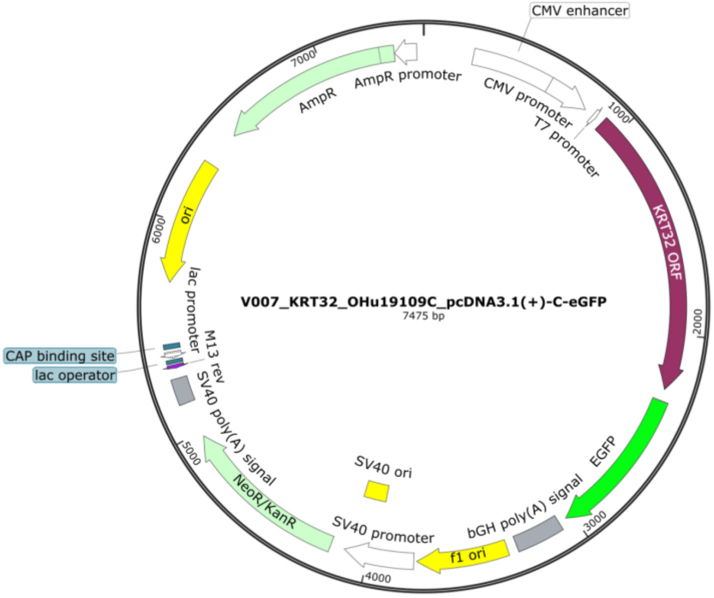


- GFP-KRT32^T99I^

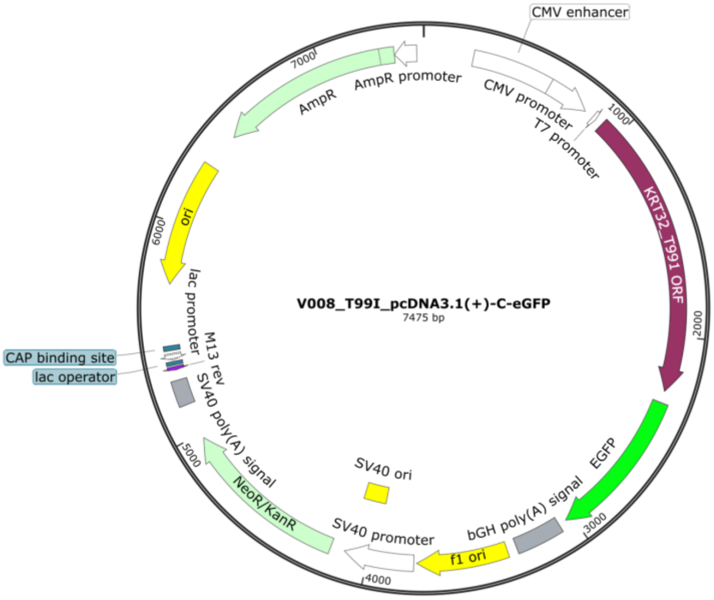

- RFP-KRT82$^{WT}$

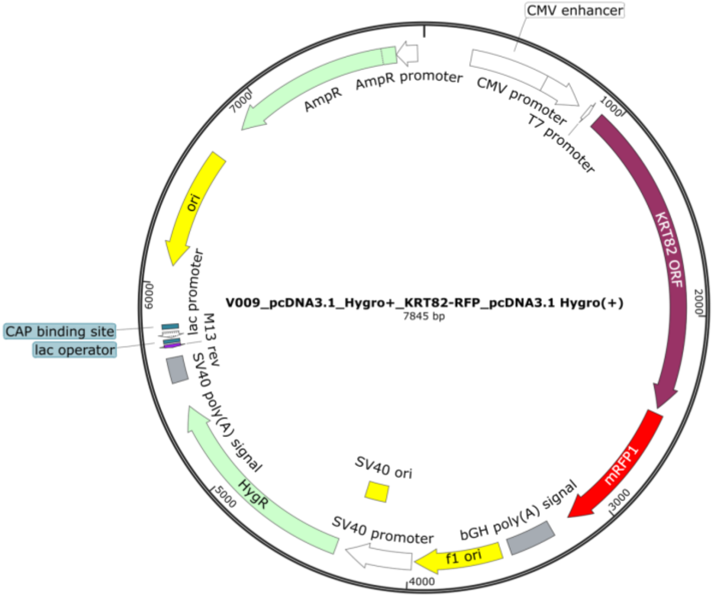


***Fluorescent signal boosters:*** (VHH/Nanobody conjugated to fluorophore)

- ChromoTek GFP-Booster Alexa Fluor 488 (Cat# gb2AF488) Proteintech Group Inc
- ChromoTek RFP-Booster Alexa Fluor 568 (Cat# rb2AF647) Proteintech Group Inc

***Post-Hoc Power Analysis:*** Continuous Endpoint, Two Independent Sample Study

| ***Number of Segment*** | |
|---|---|
| ***Mean, WT*** | 14653.54545 |
| ***Mean, MUT*** | 10858 |
| ***Subjects, WT*** | 44 |
| ***Subjects, MUT*** | 48 |
| ***Alpha*** | 0.05 |
| ***Post-hoc Power*** | 76% |

| ***Average Filament Brightness*** | |
|---|---|
| ***Mean, WT*** | 272.4726288 |
| ***Mean, MUT*** | 229.639127 |
| ***Subjects, WT*** | 44 |
| ***Subjects, MUT*** | 48 |
| ***Alpha*** | 0.05 |
| ***Post-hoc Power*** | 70% |

### ***Statistical Analysis of Metrics:***

| ***Number of Segments*** | | |
|---|---|---|
| | *Unpaired t-Test* | |
| | **WT** | **MUT** |
| ***Mean*** | 14654 | 10858 |
| ***Observations*** | 44 | 48 |
| ***Means diff. (MUT - WT) ± SEM*** | -3795 ± 1399 | |
| ***P value*** | 0.0080 | |

| ***Average Filament Brightness*** | | |
|---|---|---|
| | *Unpaired t-Test* | |
| | **WT** | **MUT** |
| ***Mean*** | 272.5 | 229.6 |
| ***Observations*** | 44 | 48 |
| ***Means diff. (MUT - WT) ± SEM*** | -42.83 ± 17 | |
| ***P value*** | 0.0151 | |

***GFP-KRT32$^{WT}$ and RFP-KRT82$^{WT}$ cotransfection (HaCaT) fluorescent microscopy images:***

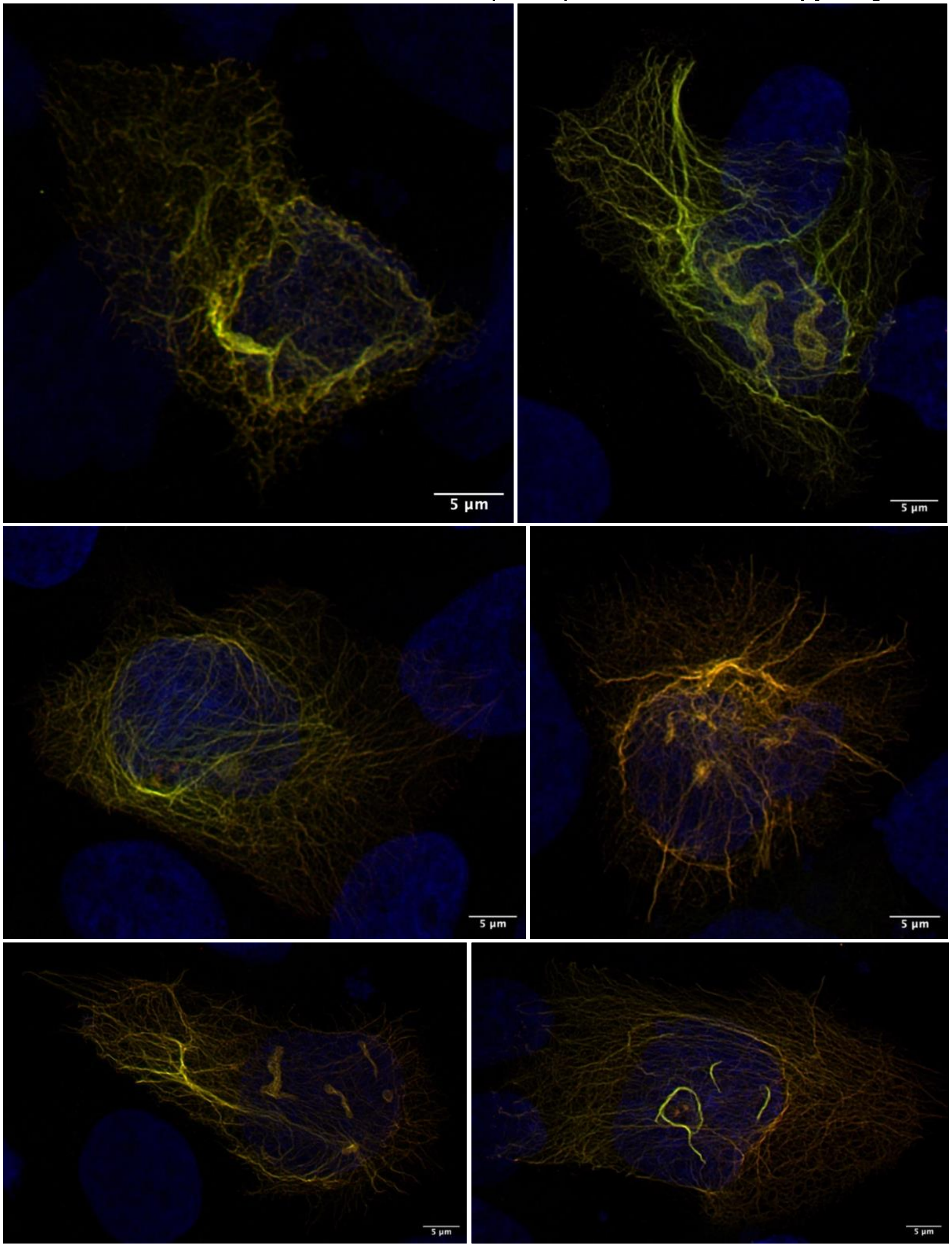

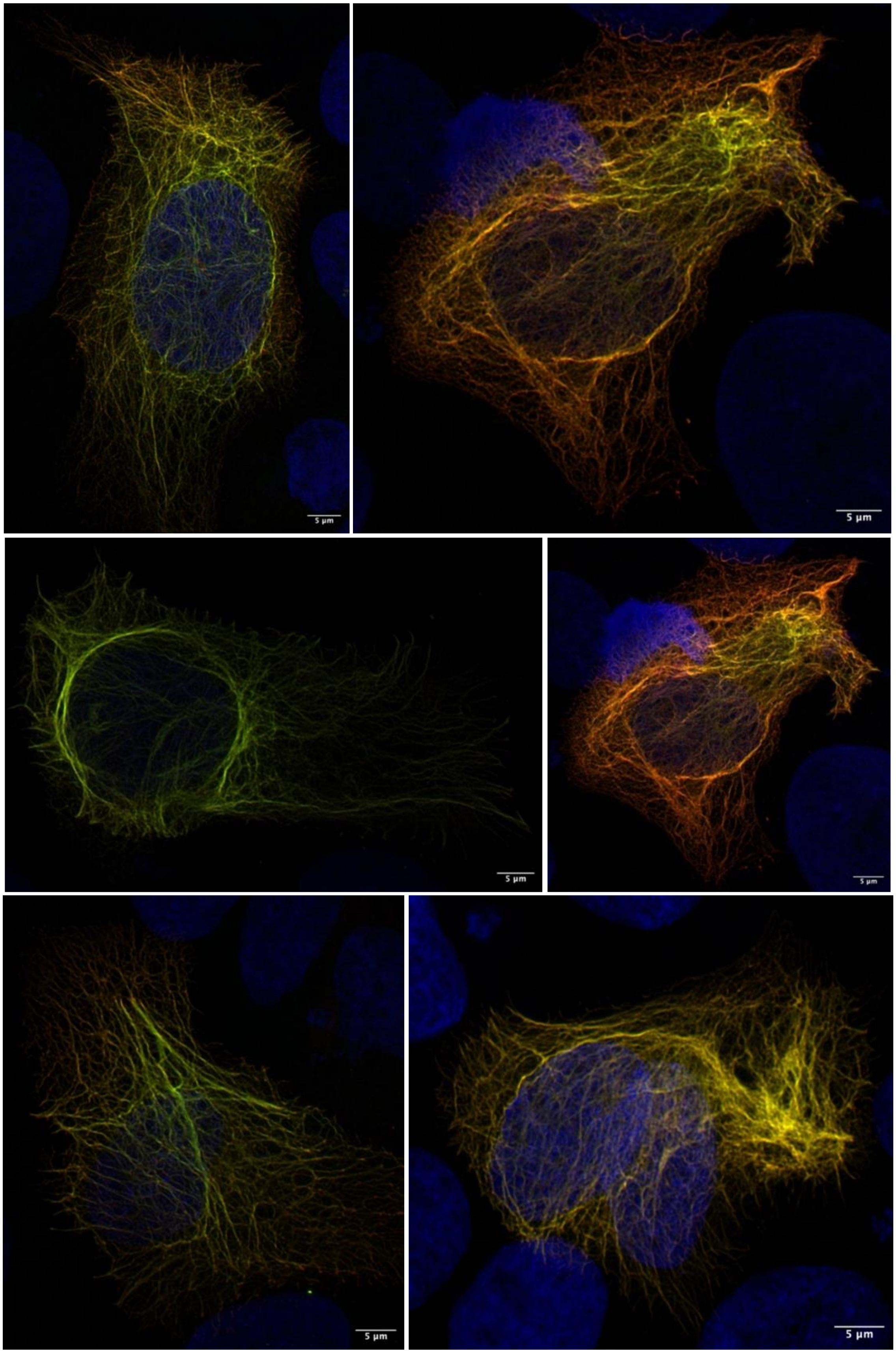
5 µm
5 µm
5 µm
5 µm
5 µm
5 µm

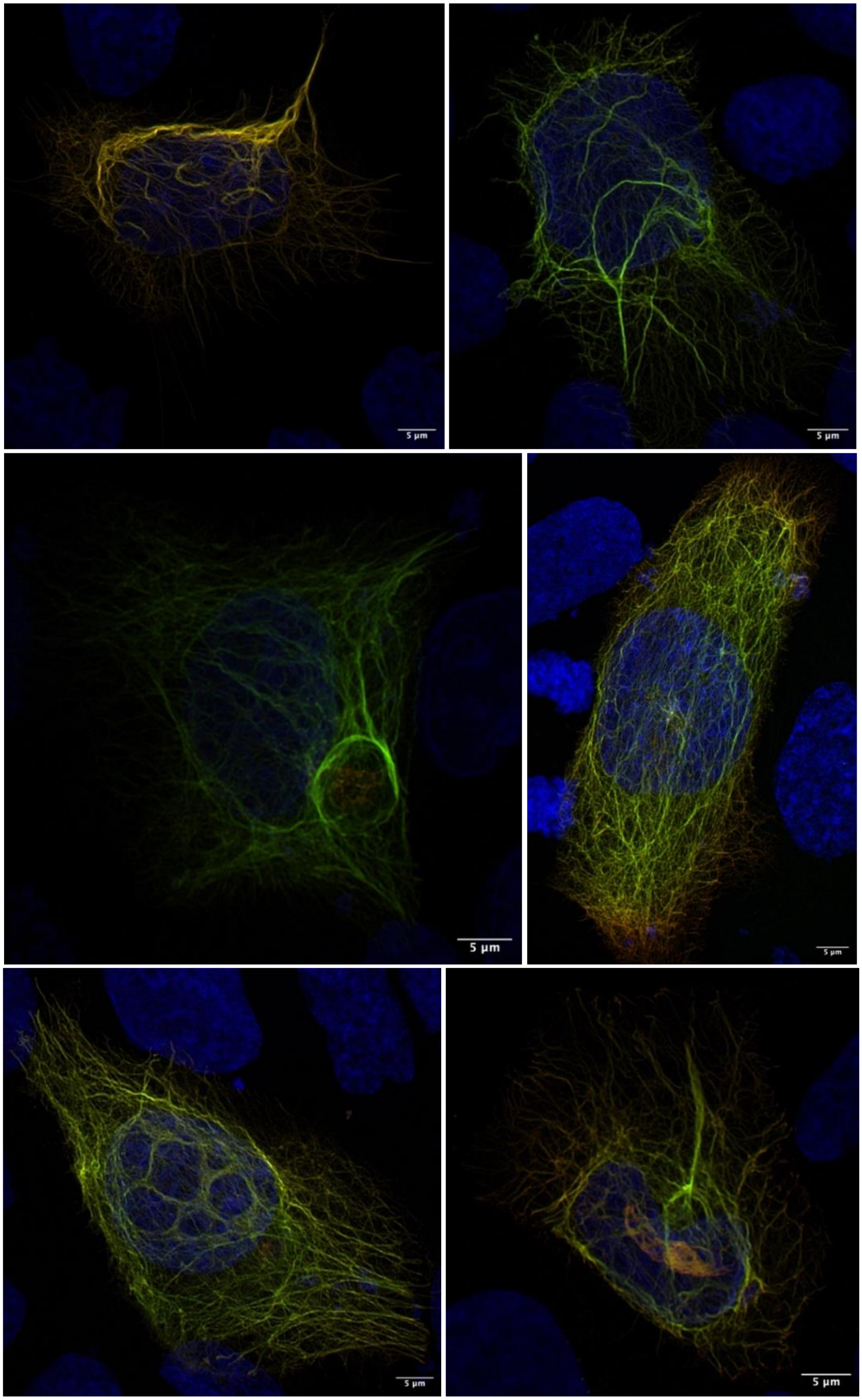
5 μm
5 μm
5 μm
5 μm
5 μm
5 μm

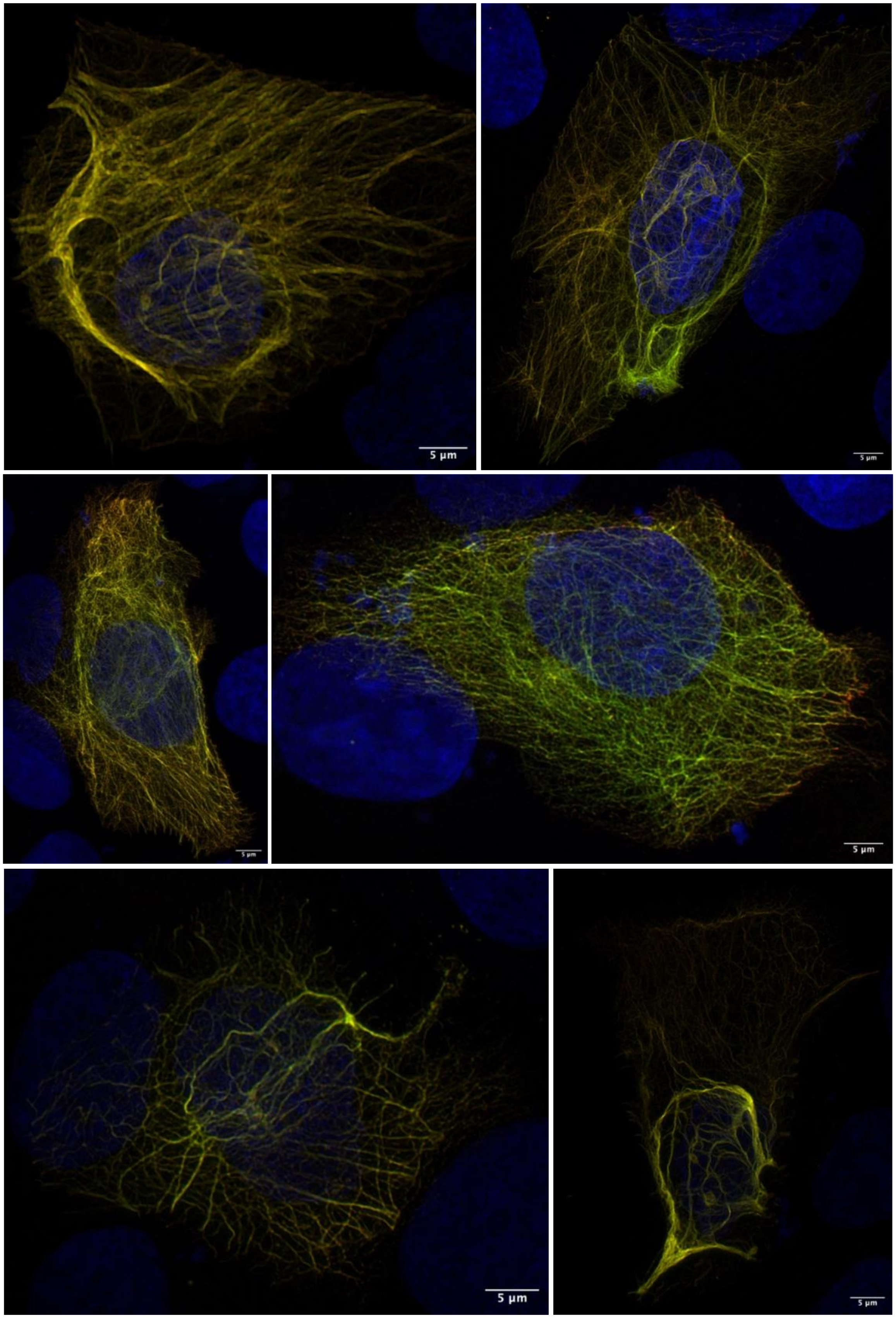
5 µm
5 µm
5 µm
5 µm
5 µm
5 µm

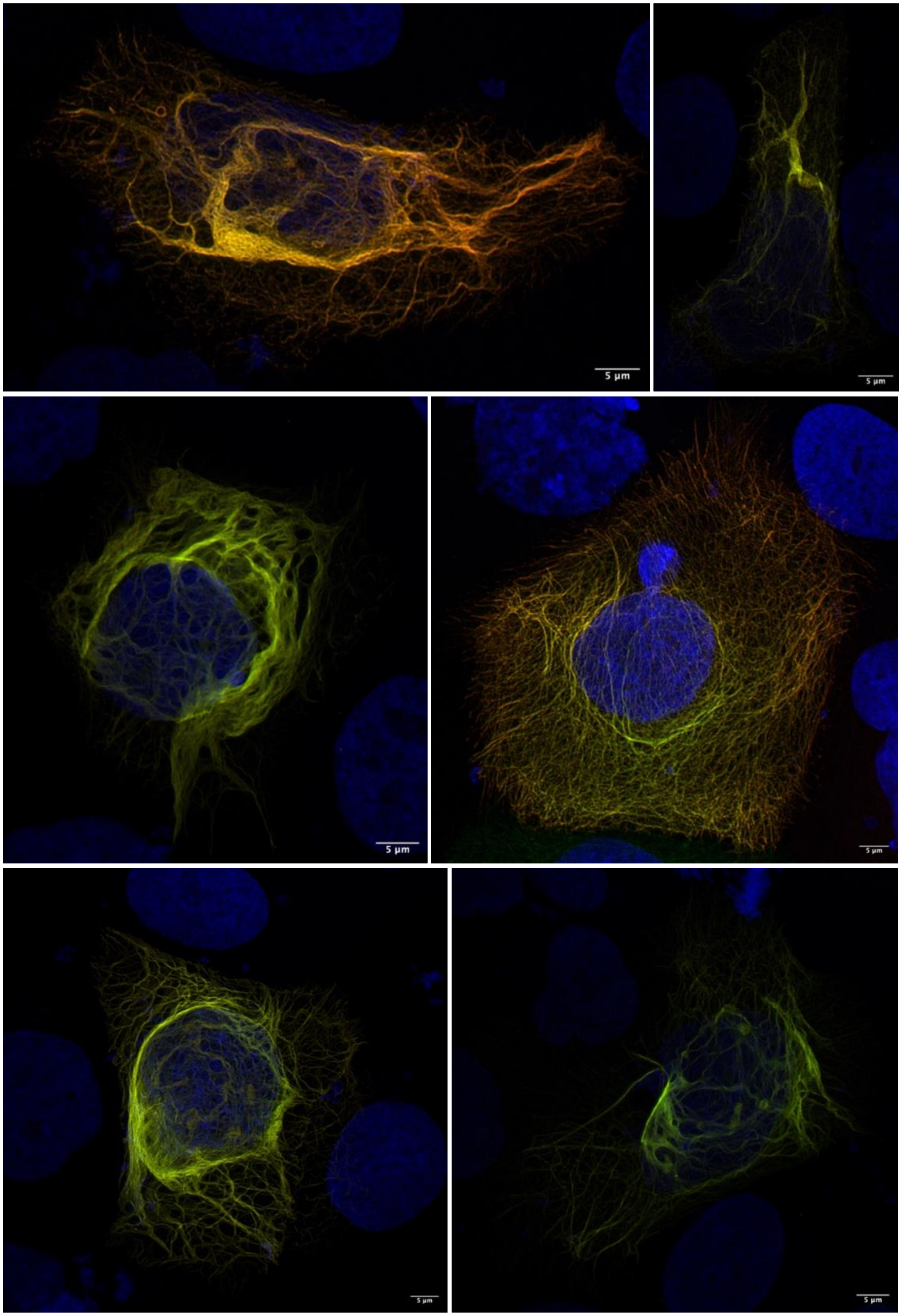
5 µm
5 µm
5 µm
5 µm
5 µm
5 µm

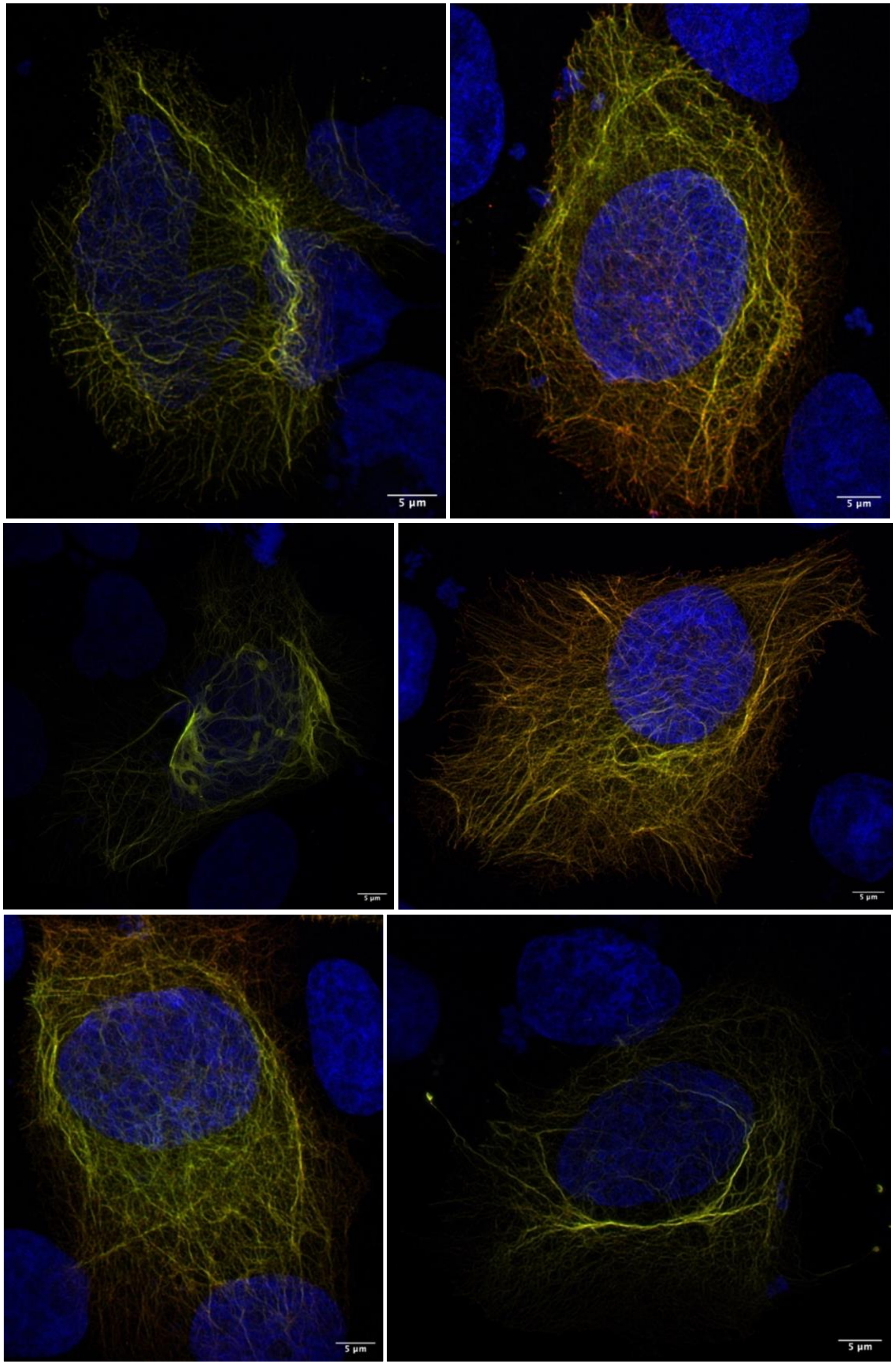
5 μm
5 μm
5 μm
5 μm
5 μm
5 μm

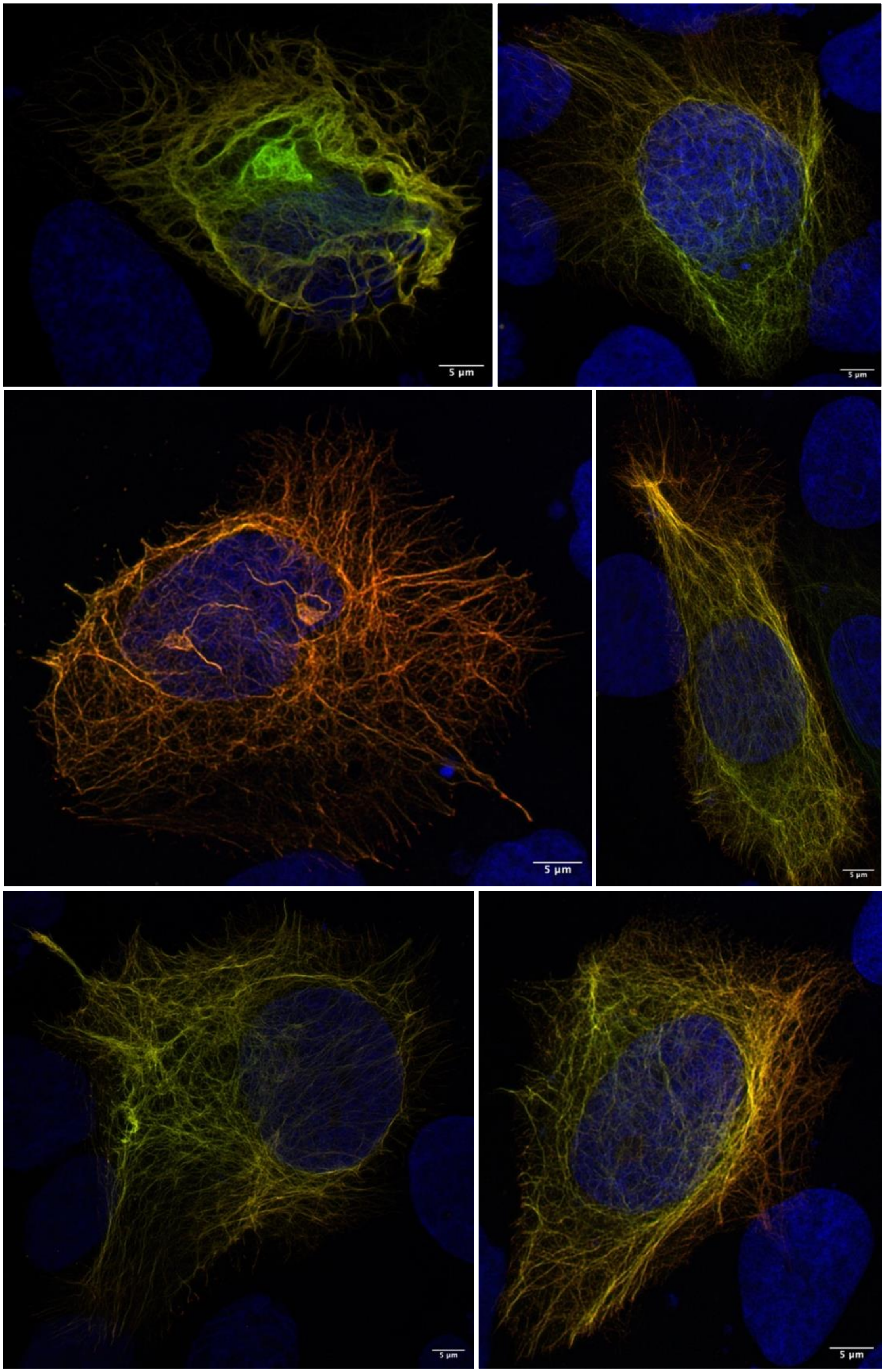
5 µm
5 µm
5 µm

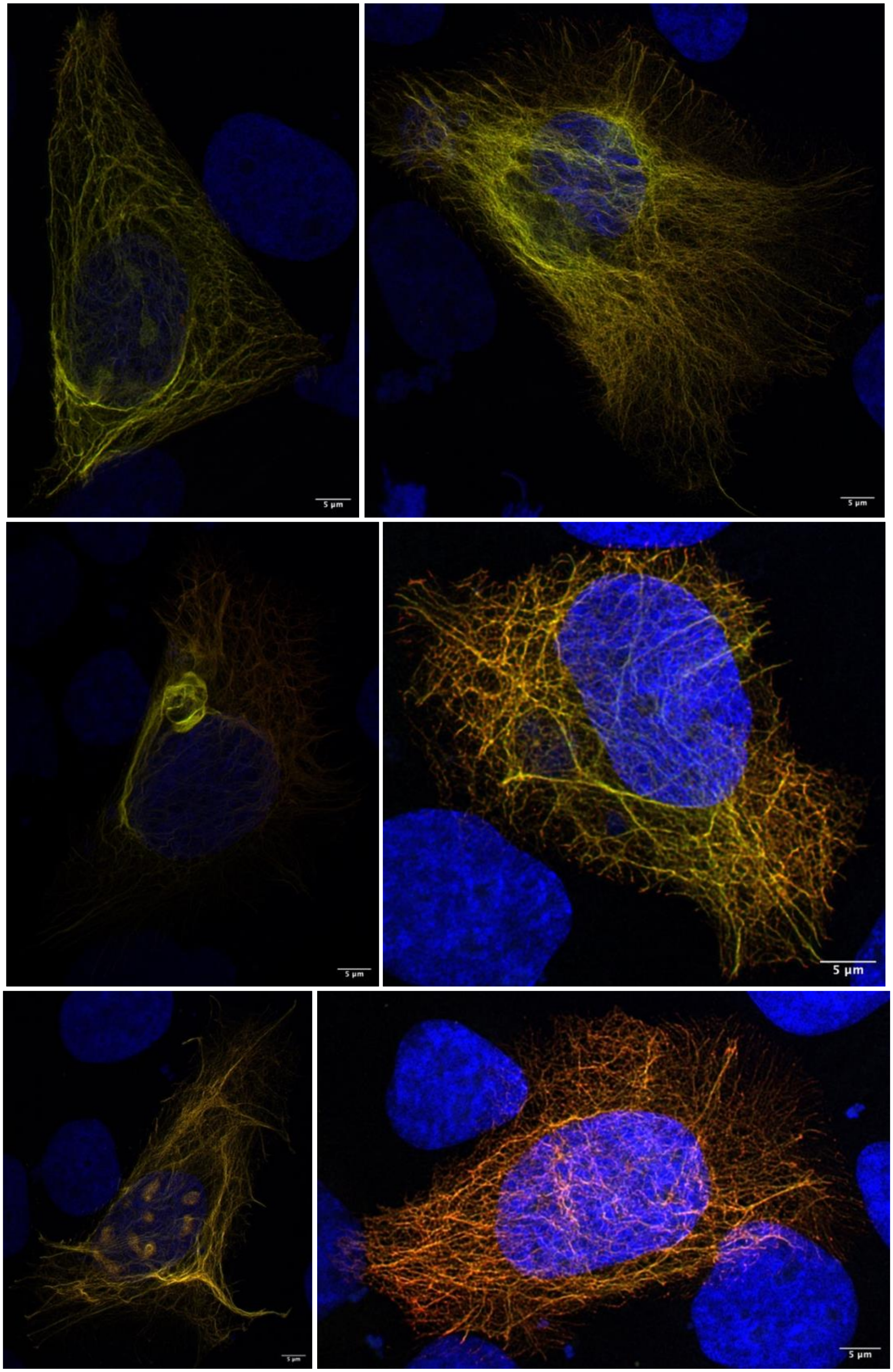
5 µm
5 µm
5 µm
5 µm
5 µm
5 µm

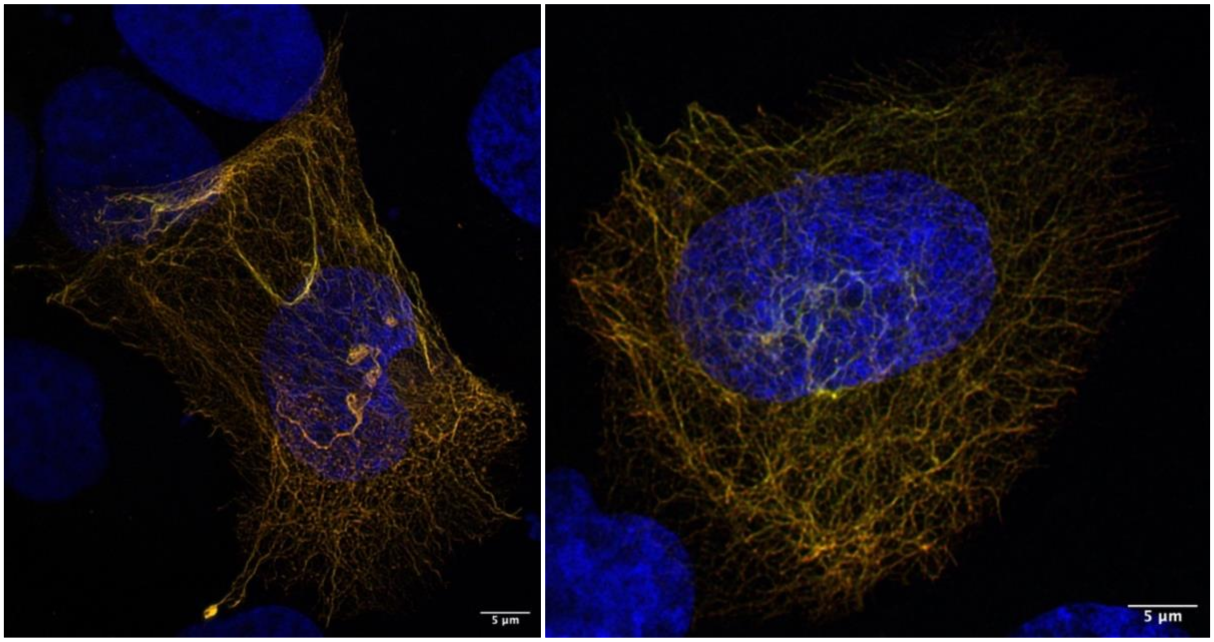
5 µm
5 µm

***GFP-KRT32$^{T99I}$ and RFP-KRT82$^{WT}$ cotransfection (HaCaT) fluorescent microscopy images:***

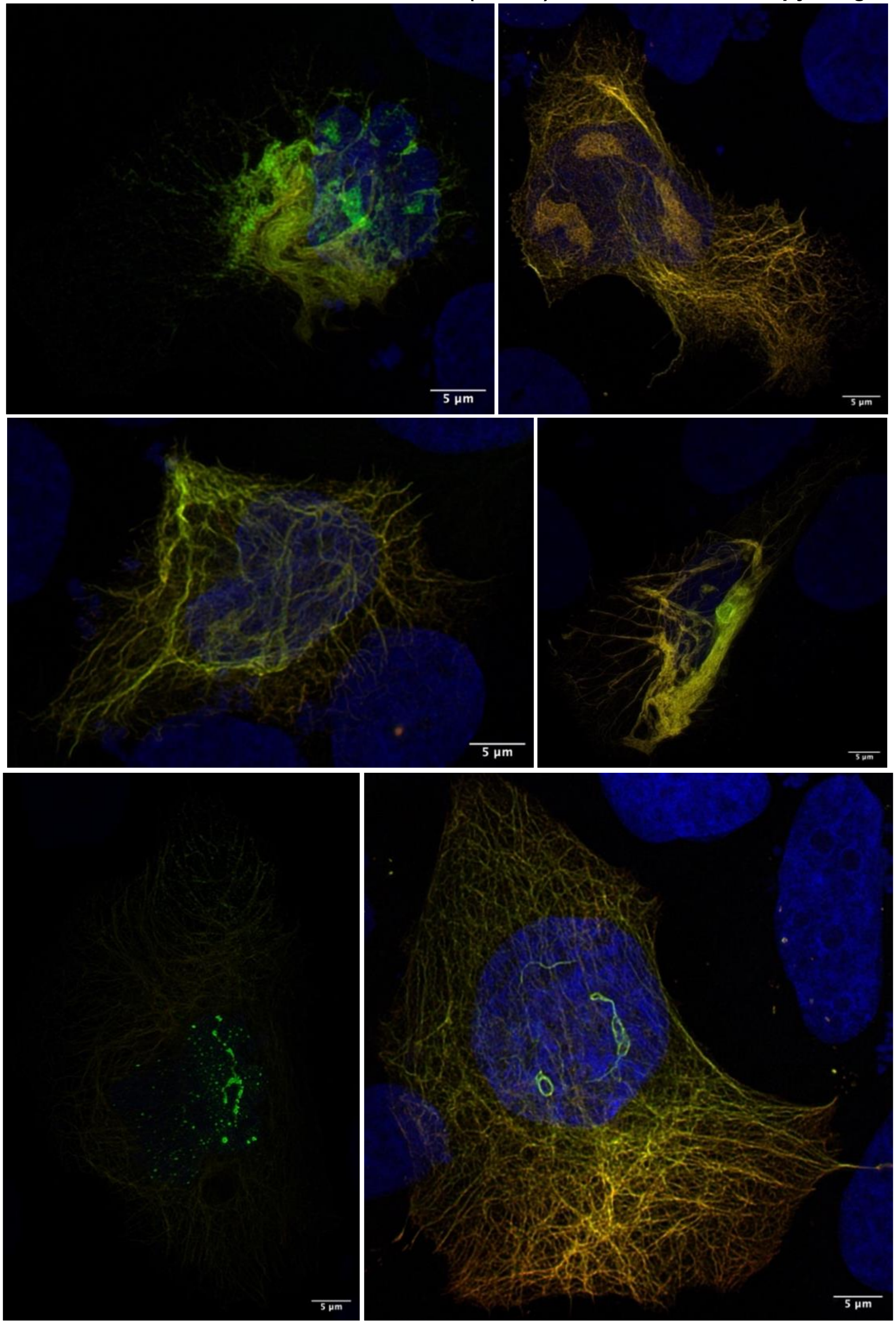

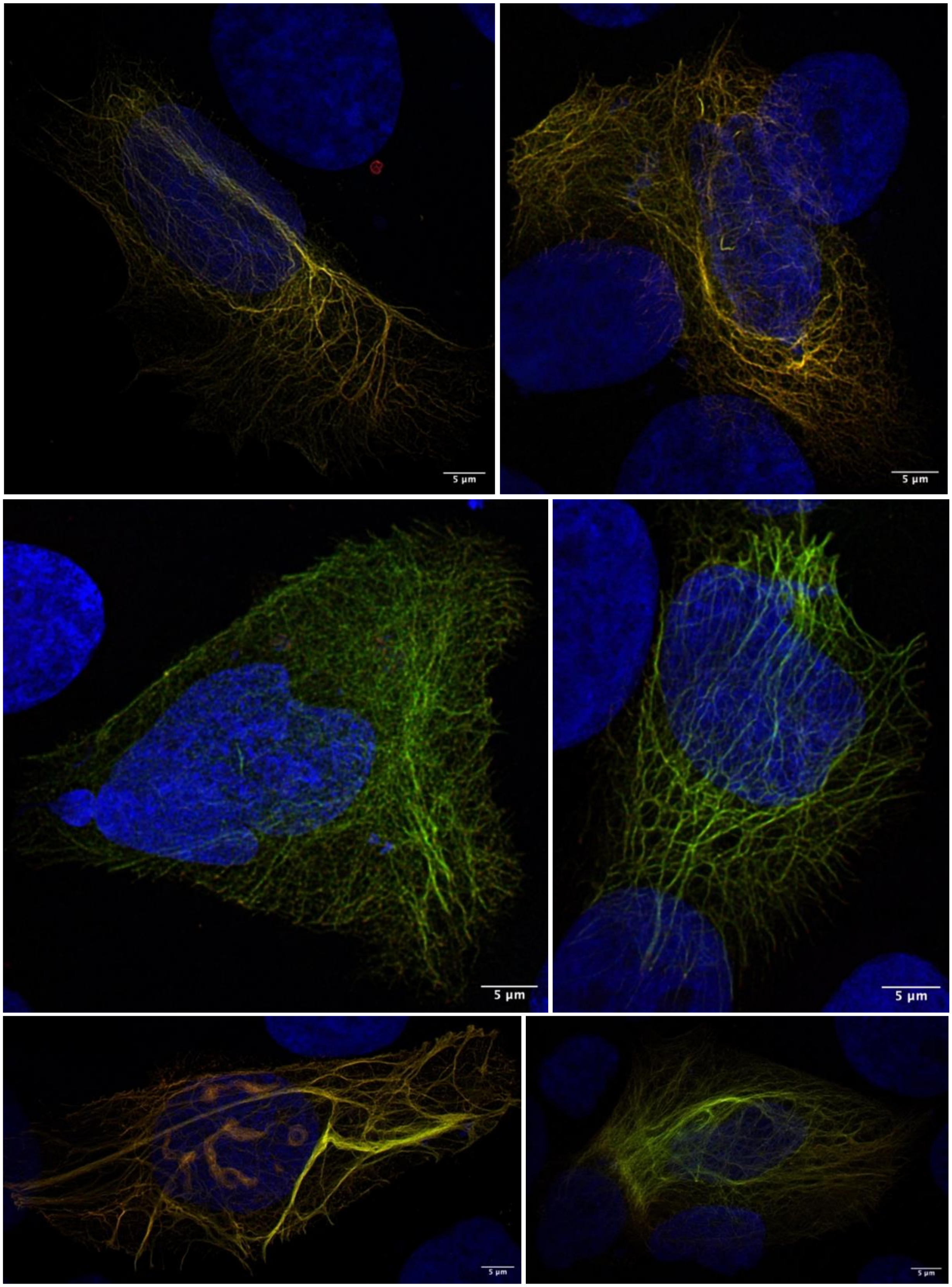
5 µm
5 µm
5 µm
5 µm
5 µm
5 µm

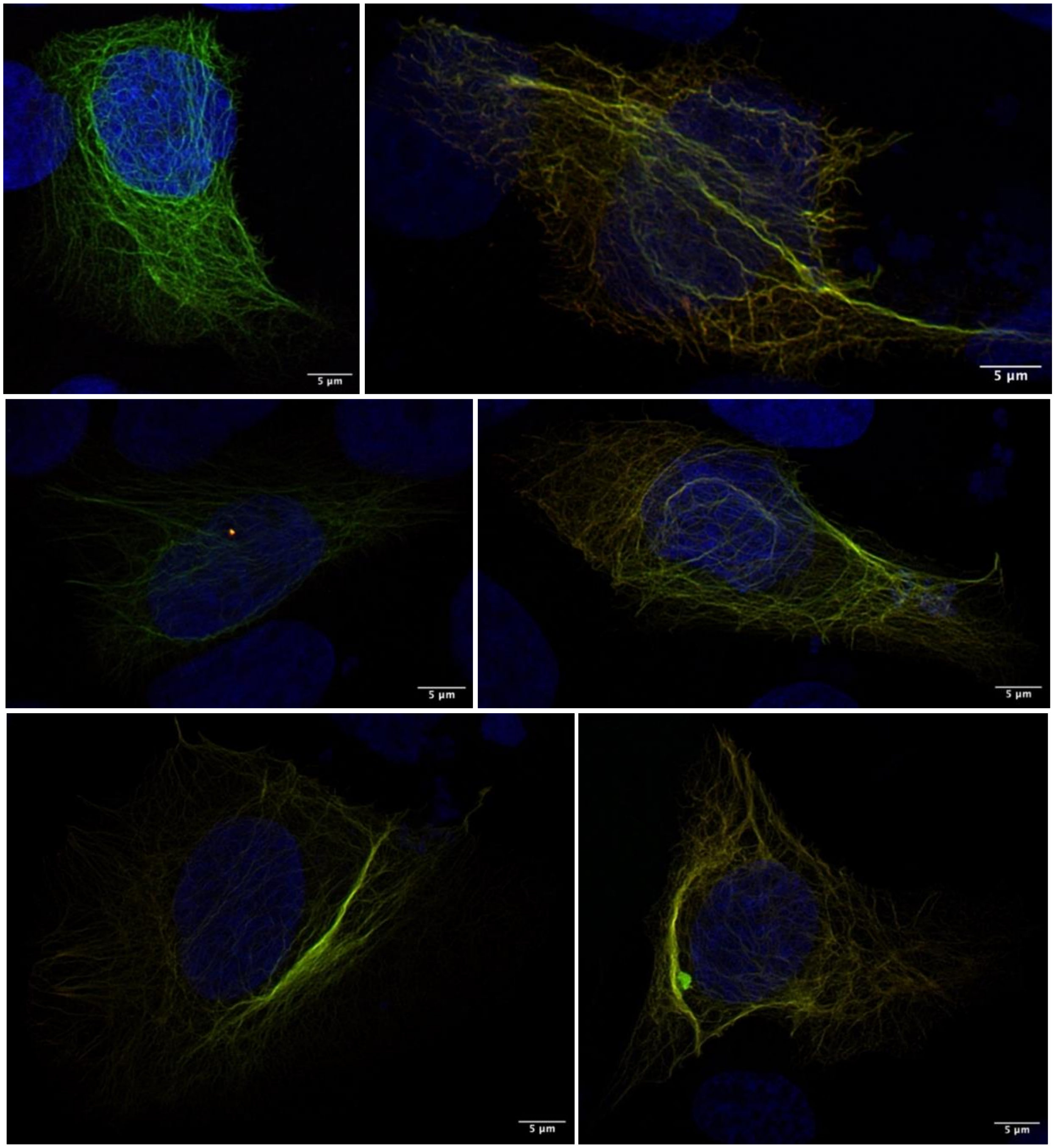
5 µm
5 µm
5 µm
5 µm
5 µm
5 µm

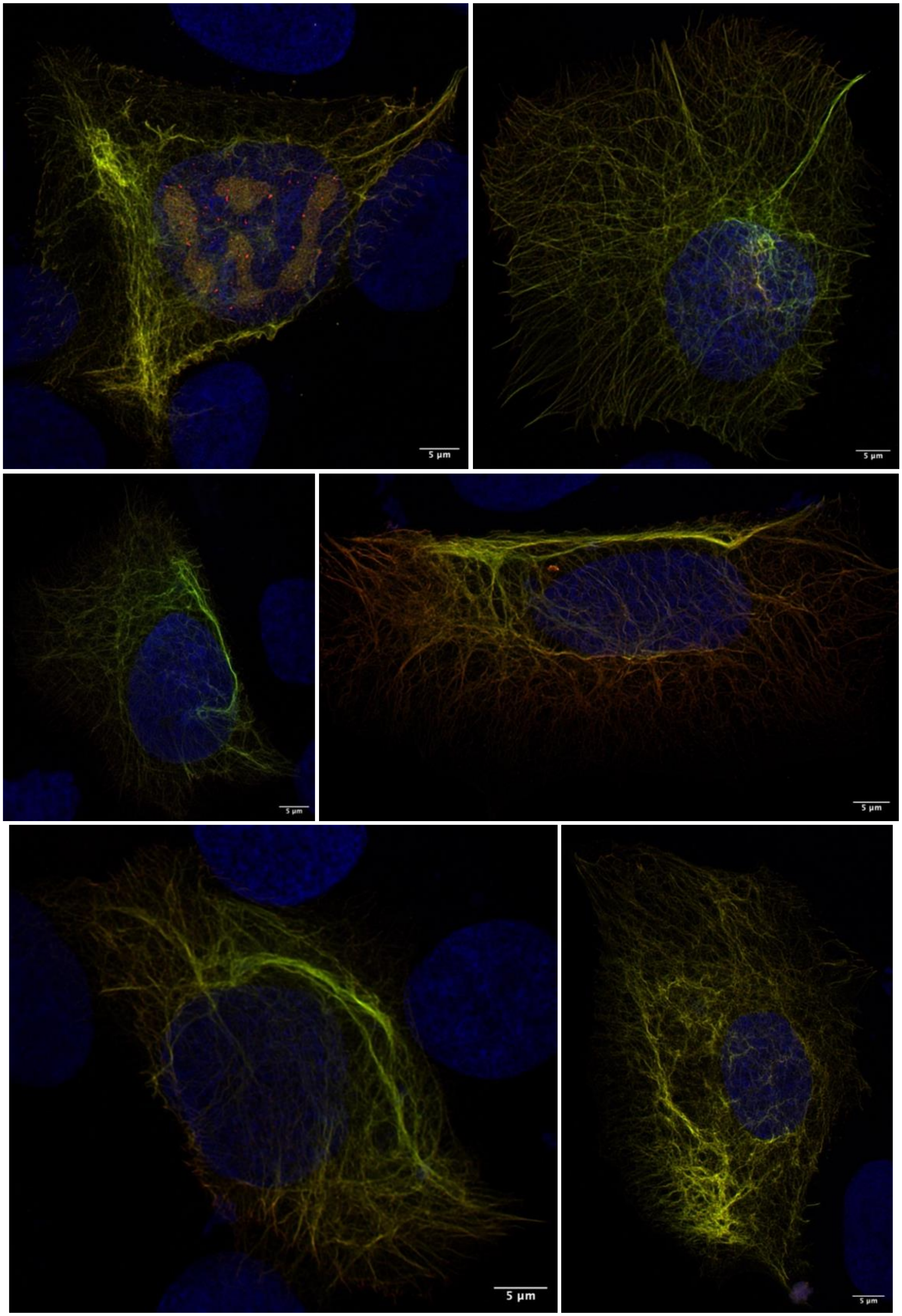
5 µm
5 µm
5 µm
5 µm
5 µm
5 µm

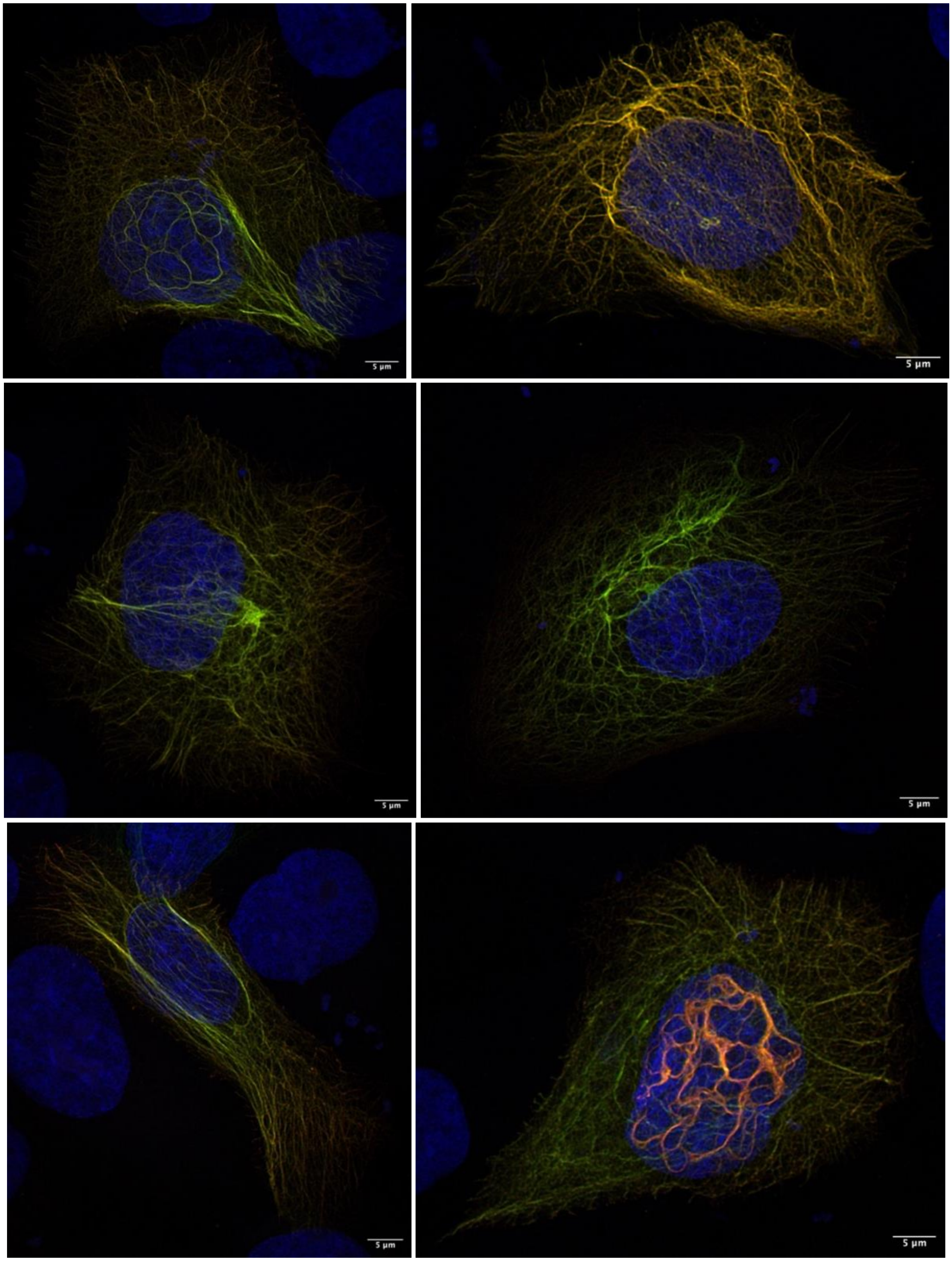
5 µm
5 µm
5 µm
5 µm
5 µm
5 µm

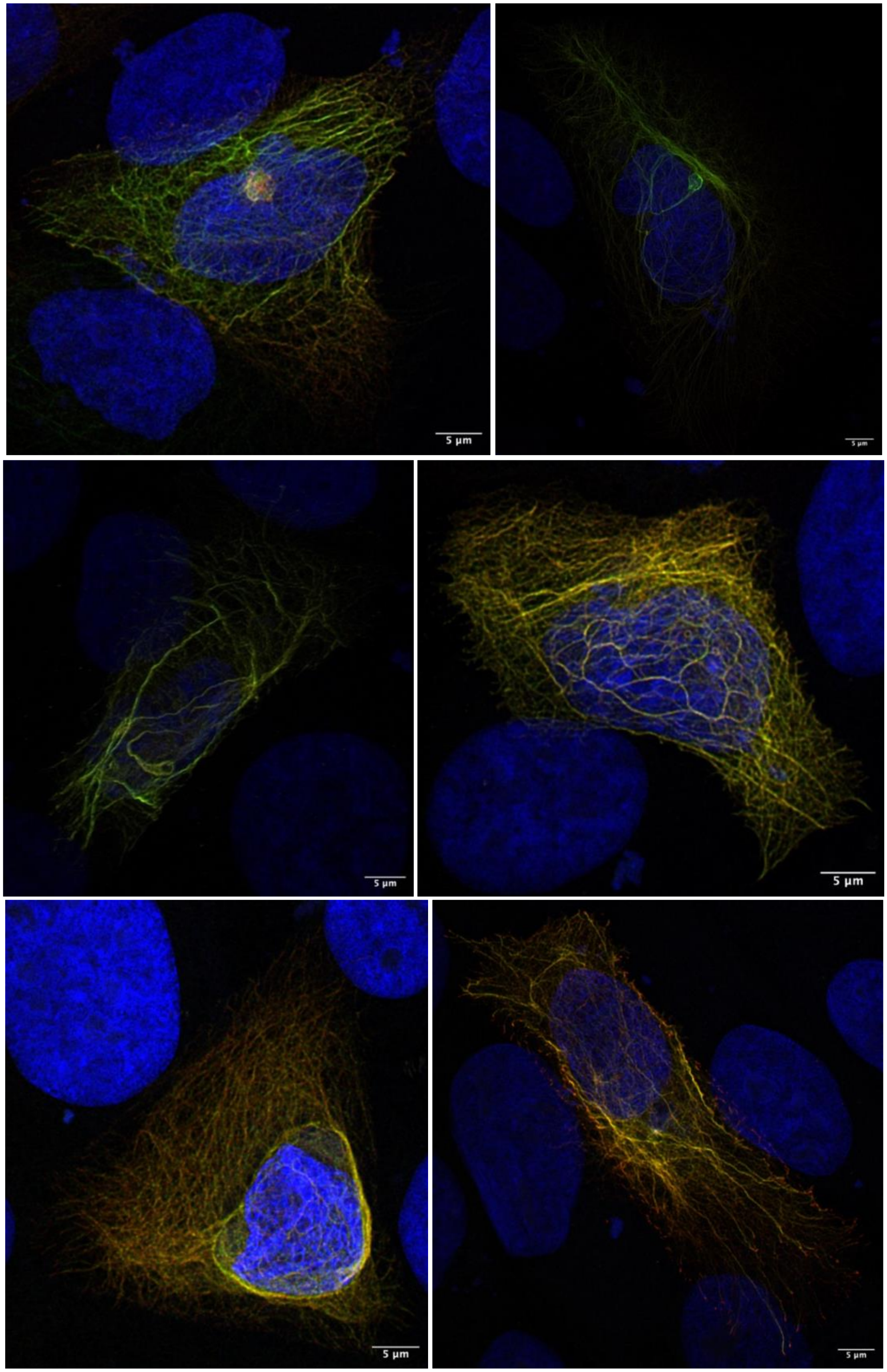
5 µm
5 µm
5 µm
5 µm
5 µm
5 µm

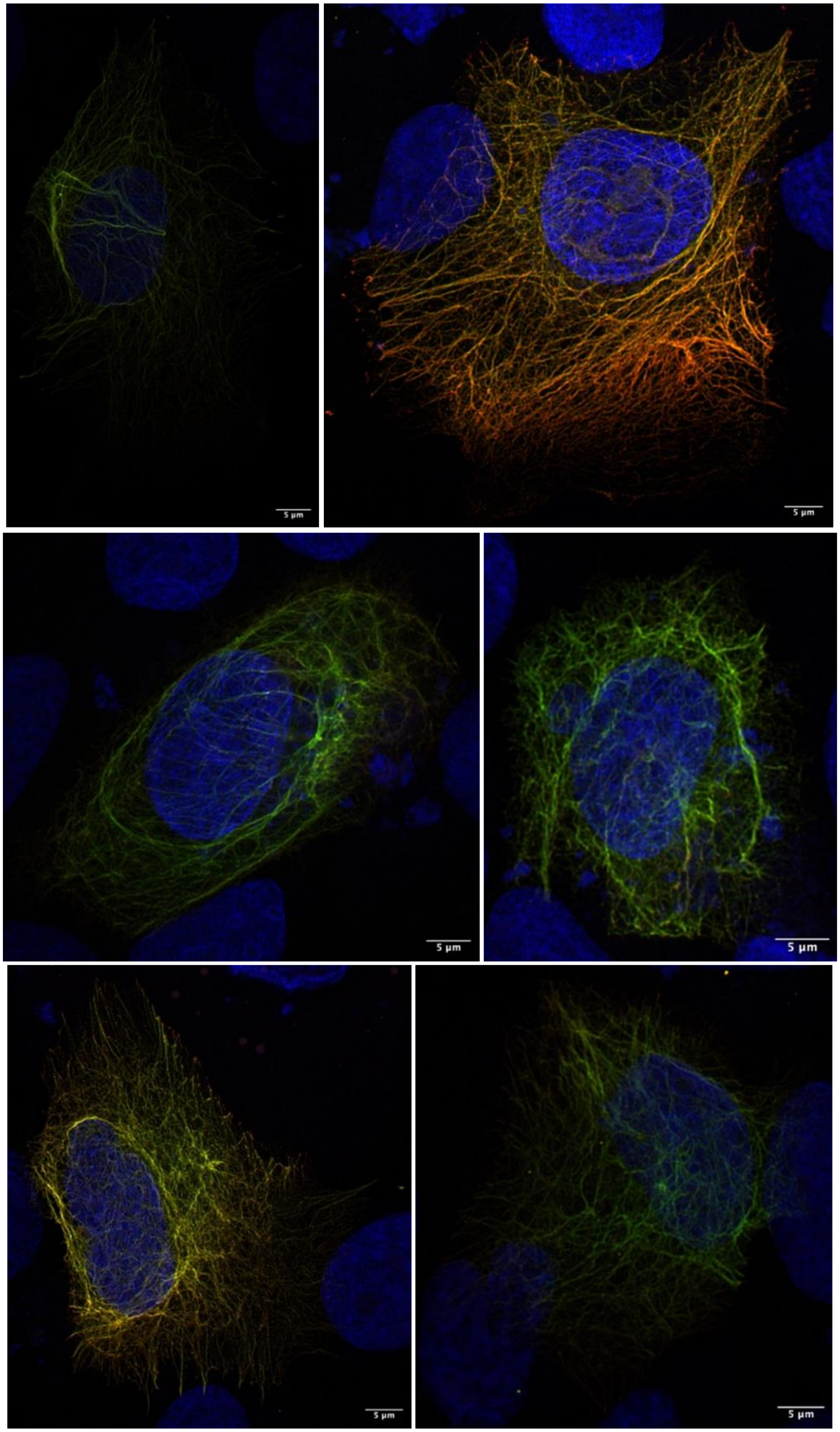
5 µm
5 µm
5 µm
5 µm
5 µm
5 µm

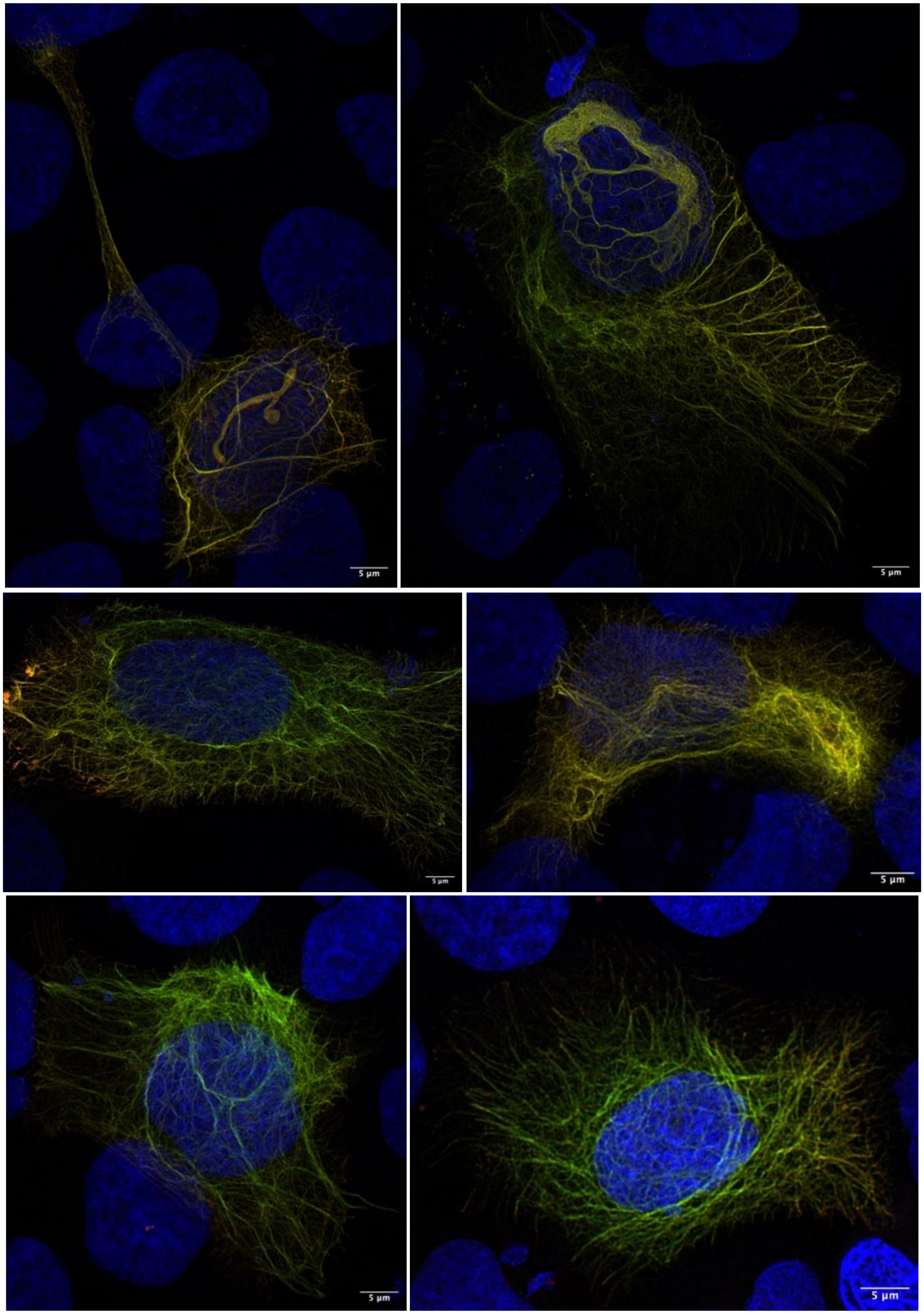
5 µm
5 µm
5 µm
5 µm
5 µm
5 µm

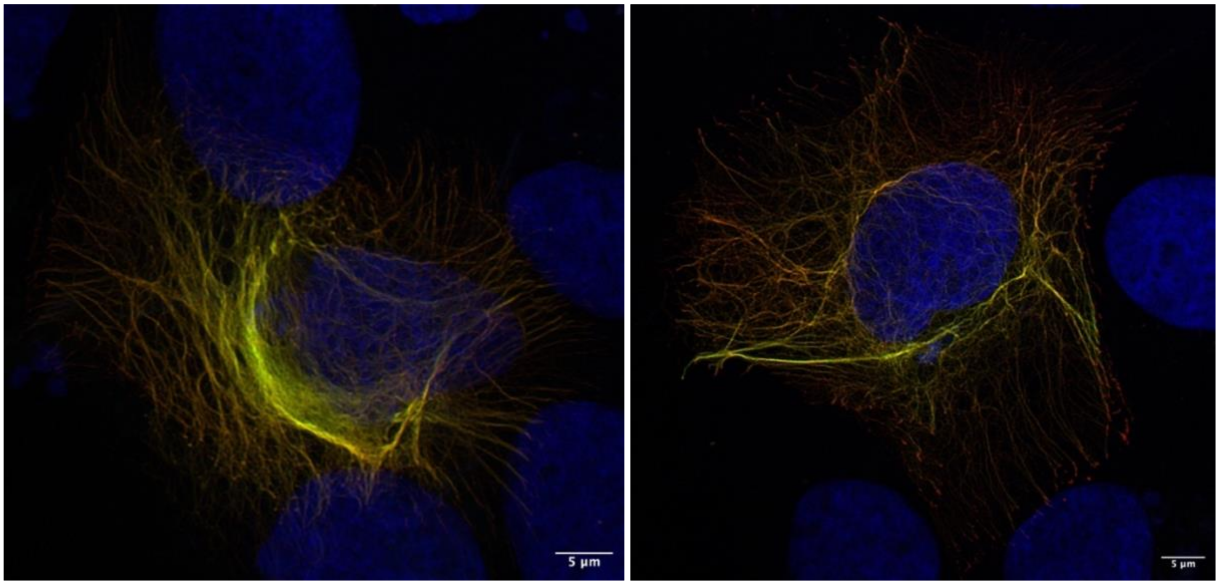
5 µm
5 µm
